# Machine learning augmented genome-wide meta-analysis of prescription opioid use in 860,000 individuals

**DOI:** 10.64898/2025.12.07.25341785

**Authors:** Lisa Eick, Laura Birgit Luitva, Kristi Krebs, Sakari Jukarainen, Sami Kulju, FinnGen, Estonian Biobank research team, Maiju Marttinen, Manuel A. Rivas, Andrea Ganna, Lili Milani, Zhiyu Yang, Tuomo Kiiskinen

## Abstract

Opioid analgesics are widely prescribed for pain, yet individuals show substantial variation in medical opioid use. To investigate the genetic basis of prescription-derived intake, we analyzed 859,675 Europeans across three biobanks. Prescription records were harmonized to cumulative oral morphine equivalents (OME), yielding three outcomes: any opioid prescription, cumulative dose among users, and population-level dose including non-users. Genome-wide meta-analyses identified 78, 20, and 135 loci, respectively (234 independent signals across 145 regions). All traits were highly correlated and strongly overlapped with pain-related genetics, though cumulative dose among users captured a more distinct dose-intensity component.

To detect deviations from expected medical use, we trained gradient-boosted models to derive early-onset and excess-dose phenotypes. Early onset showed no genome-wide associations and mirrored pain architecture. Excess dose identified a significant signal at rs58099562 in high LD with the CYP2D6*4 loss-of-function allele and correlated more with psychiatric and substance-use traits.

These results show that conventional prescription traits primarily reflect pain biology, whereas disproportionately high dosing captures distinct neuropsychiatric and pharmacokinetic liability.

**Graphical Abstract:** Phenotype definitions, meta-analysis, and machine learning-based refinement of opioid prescription traits.
(a) Three opioid prescription phenotypes were defined: binary prescription status (RxExpPop), cumulative dose among users (RxDoseUser), and combined dosage plus binary prescription (RxDosePop).
(b) Genome-wide association studies were performed across multiple biobanks and meta-analyzed using METAL.
(c) Machine learning was applied to refine overuse phenotypes, resulting in early onset (RxOverUse_Onset) and high dose (RxOverUse_Amount) subtypes.
(d) Downstream analyses included genetic correlation, gene annotation, biological interpretation, and cross-study comparisons.

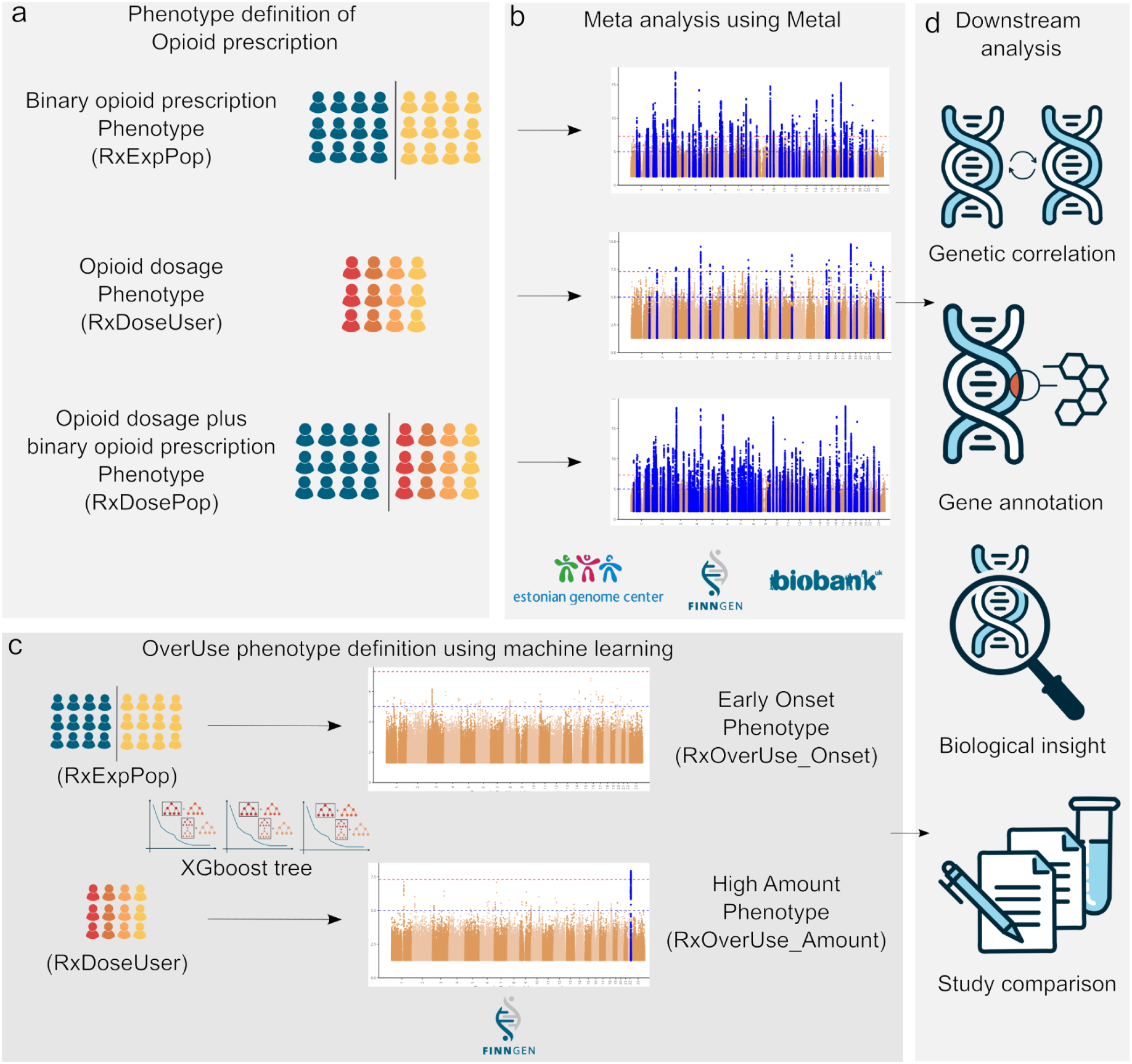

## Main

Relief of pain is foundational to medical care, and opioid analgesics remain central to analgesia when pain is acute or severe, including postoperative care, trauma, cancer pain, and palliative treatment. In these settings, and when other options fail, opioids provide rapid and reliable pain control and are therefore widely used in routine practice^1^. At the same time, lifelong opioid exposure varies enormously between individuals, ranging from never being prescribed to sustained high-dose use in severe conditions such as cancer pain^2,3^. While the neurobiology of opioids is well described, including central reward circuitry^4,5^ and respiratory and gastrointestinal effects^6,7^, the determinants of interindividual differences in prescription-derived exposure and total cumulative dose remain less well characterized genetically, as most prior work has focused on opioid use disorder (OUD) rather than prescribing patterns^8^.

The need for effective analgesia is immense. Chronic pain, defined as pain lasting more than three months^9^, is among the most common and disabling health conditions worldwide. Global Burden of Disease analyses rank pain disorders among the leading causes of years lived with disability: low back pain and migraine are first and second globally, and neck pain is within the top ten^10^. Recurrent tension-type headache alone affects about 1.9 billion people, making it the most prevalent symptomatic chronic condition^9^. This burden underscores the necessity of potent analgesics.

Non-opioid treatments, such as nonsteroidal anti-inflammatory drugs, anticonvulsants, antidepressants, and gabapentinoids, offer limited and often short-term relief for pain. Consequently, opioids remain essential for managing acute, postoperative, cancer-related, and palliative pain^1,11^.

However, opioids carry substantial risks. Even when prescribed appropriately, opioids can cause sedation, gastrointestinal complications, and respiratory depression^6,7^. Their direct effects on the mesolimbic dopamine system confer reinforcing potential, and the risk of prolonged use and misuse rises with longer duration and higher doses, motivating caution in ongoing therapy^4,5^. Against this backdrop, understanding why some patients accumulate far higher total cumulative doses than others, including the contribution of genetic factors, is a pressing question^12^.

The liberalization of opioid prescribing in the late 1990s catalyzed a public-health crisis: prescription opioid use expanded rapidly, OUD rates rose, and synthetic opioids became the leading driver of drug-related deaths^13,14^. These events sharpen the need to disentangle the pathways that shape prescription-derived exposure and dose at the population level, pain biology and indication, multimorbidity and healthcare utilization, psychiatric/substance-use liability, and pharmacokinetics, and to determine how genetics contributes to interindividual differences along these pathways^12^.

Given the central role of opioids in pain management, understanding the genetic basis of prescription-derived opioid use and dosage is an important research goal. Previous GWAS have focused mainly on opioid use disorder (OUD) case–control status, identifying variants in genes such as OPRM1^8^, and on genetic factors influencing opioid metabolism and dosing, which are also used to inform prescription recommendations, including variants in CYP2D6, OPRM1, and COMT^15^. Clarifying these genetic influences is important for understanding the biological pathways involved in pain treatment and for identifying individuals who may require unusually high or low opioid doses.

Here we perform a cross-biobank GWAS of prescription-derived opioid exposure and total cumulative dose in a combined sample of 859,675 participants from FinnGen, Estonian Biobank (EstBB), and UK Biobank (UKBB). Our framework separates exposure from dose intensity and introduces a machine learning model-adjusted overuse phenotypes that capture deviation from expected use given an individual’s medical history. To make intake comparable across compounds and routes, we harmonized every opioid prescription to oral morphine equivalents (OME). We test the hypotheses that exposure-oriented signals primarily index pain/indication biology and multimorbidity, whereas dosage-deviation is enriched for psychiatric/substance-use liability and pharmacokinetic pathways.

## Results

### Opioid prescription in three biobanks: study design and baseline clinical associations

We analyzed opioid prescriptions among 859,675 participants from FinnGen, Estonian Biobank (EstBB), and UK Biobank (UKBB), of whom 401,764 had at least one prescription. Prescription data were harmonized into total oral morphine equivalents (OME) to assess three traits: prescription exposure (RxExpPop; ever vs never), cumulative dose among users (RxDoseUsers; log_n_ total cumulative OME), and population-level dose (RxDosePop; log_n_ total cumulative OME across the full cohort with non-users set to zero). Prescription patterns were broadly consistent across cohorts, with strong correlations in prescribed opioids (r=0.71–0.77, Supplementary Fig. 1). In FinnGen, opioid use correlated positively with older age, higher BMI, pain-related conditions, mental health disorders, cancer, and smoking history, highlighting a multifactorial clinical profile. Complete association statistics are presented in Supplementary Tables 1-6 and Supplementary Figures 2-3.

### Meta-analysis of GWAS across three biobanks

We performed genome-wide association studies (GWAS) separately in FinnGen, Estonian Biobank (EstBB), and UK Biobank (UKBB), finding high genetic consistency across all three opioid prescription phenotypes (binary exposure, dose among users, and population dose). Genetic correlations were particularly strong between matched phenotypes across biobanks, typically exceeding 0.8, demonstrating robust reproducibility despite differing healthcare settings and prescription capture methods.

Leveraging the strong cross-cohort consistency, we conducted fixed-effects meta-analyses across the three biobanks, substantially increasing statistical power and identifying a total of 234 independent genome-wide significant variants mapping to 145 unique genomic loci (Table 2). The largest number of associations was observed for the population-level dose phenotype (RxDosePop; 135 loci), reflecting its enhanced power from integrating both exposure and dose information. Prescription exposure (RxExpPop; 78 loci) and dose among users (RxDoseUsers; 20 loci) were less powerful but still yielded multiple significant associations.

**Table 2.**
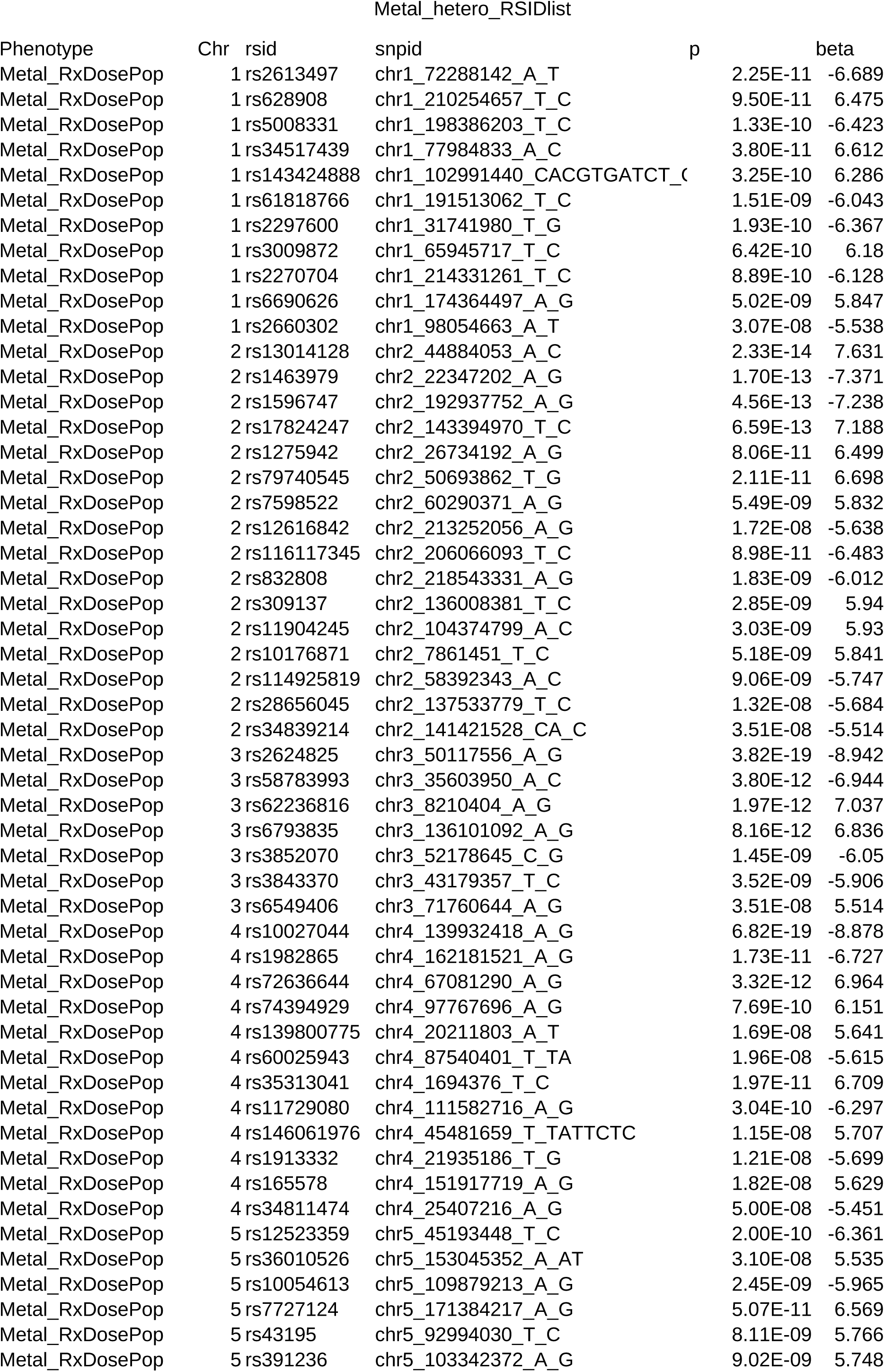
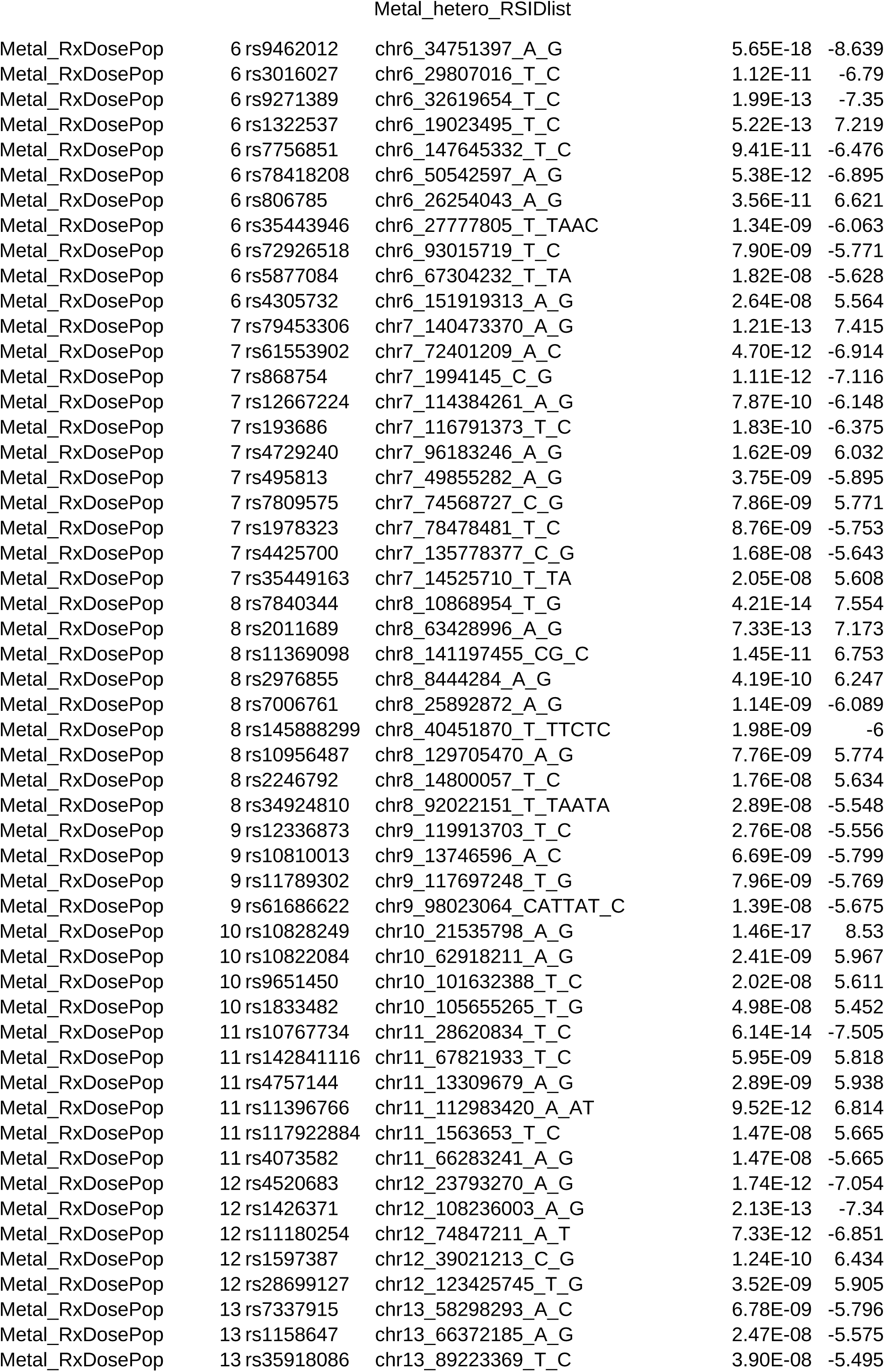
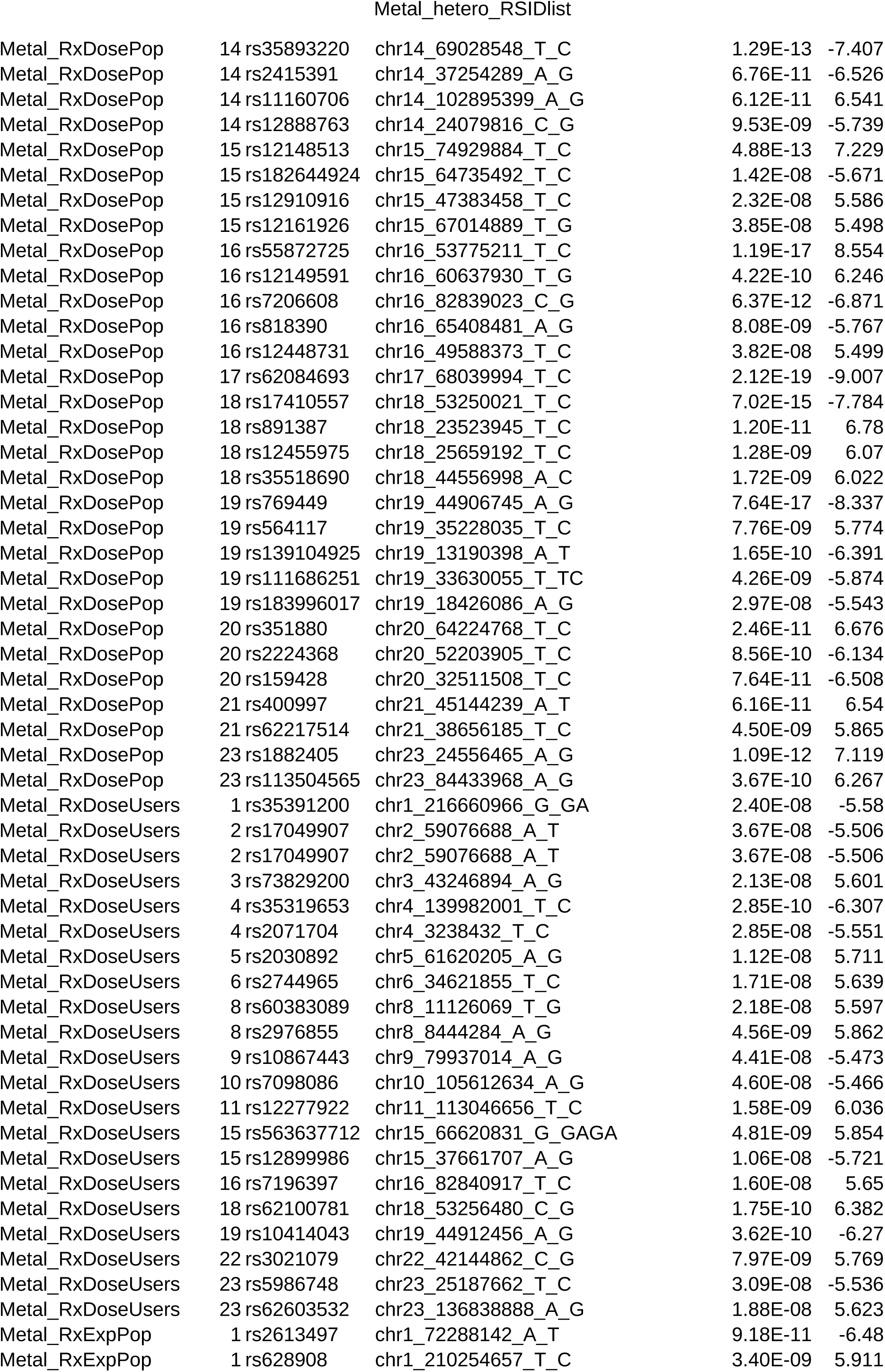
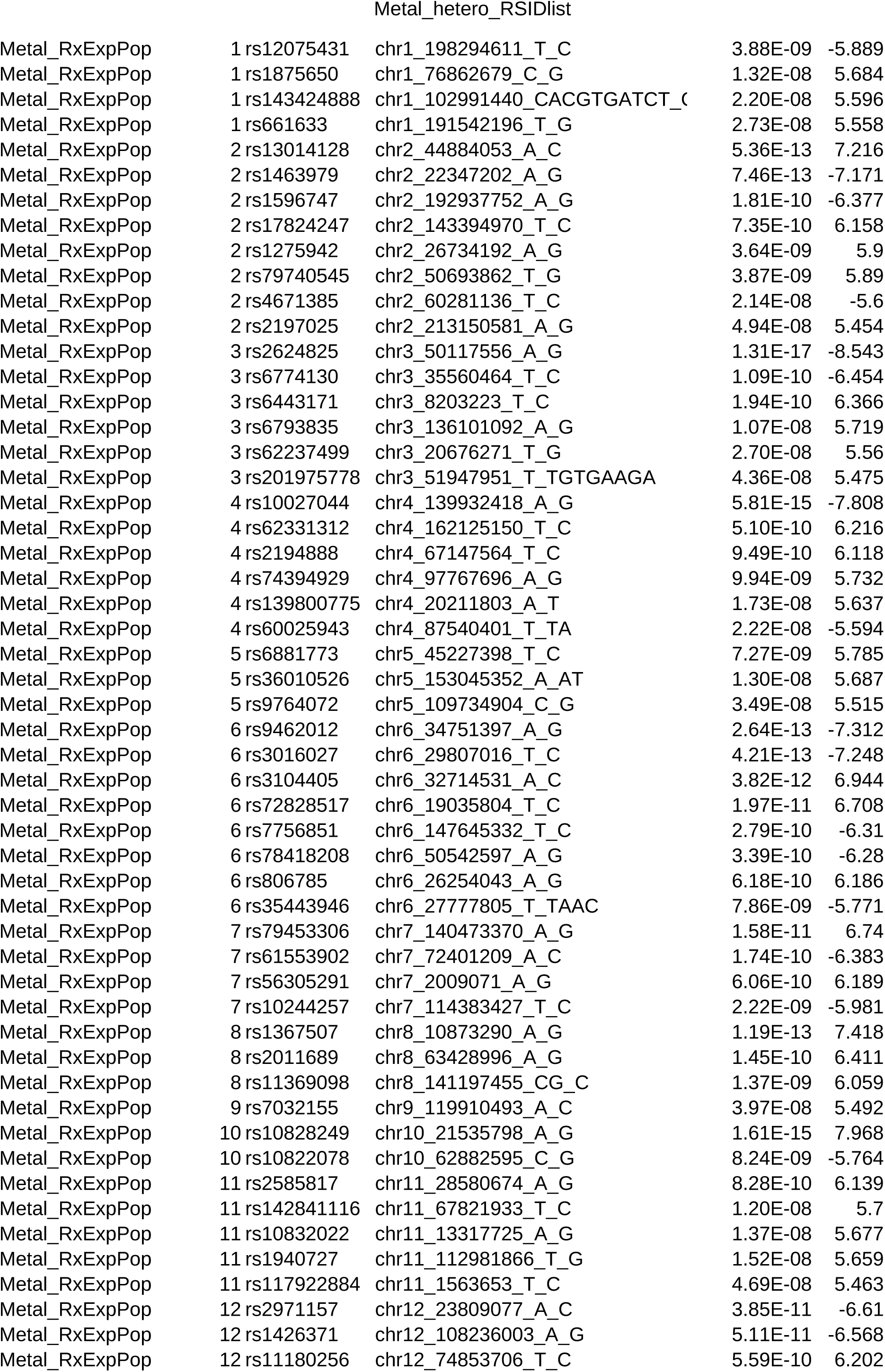
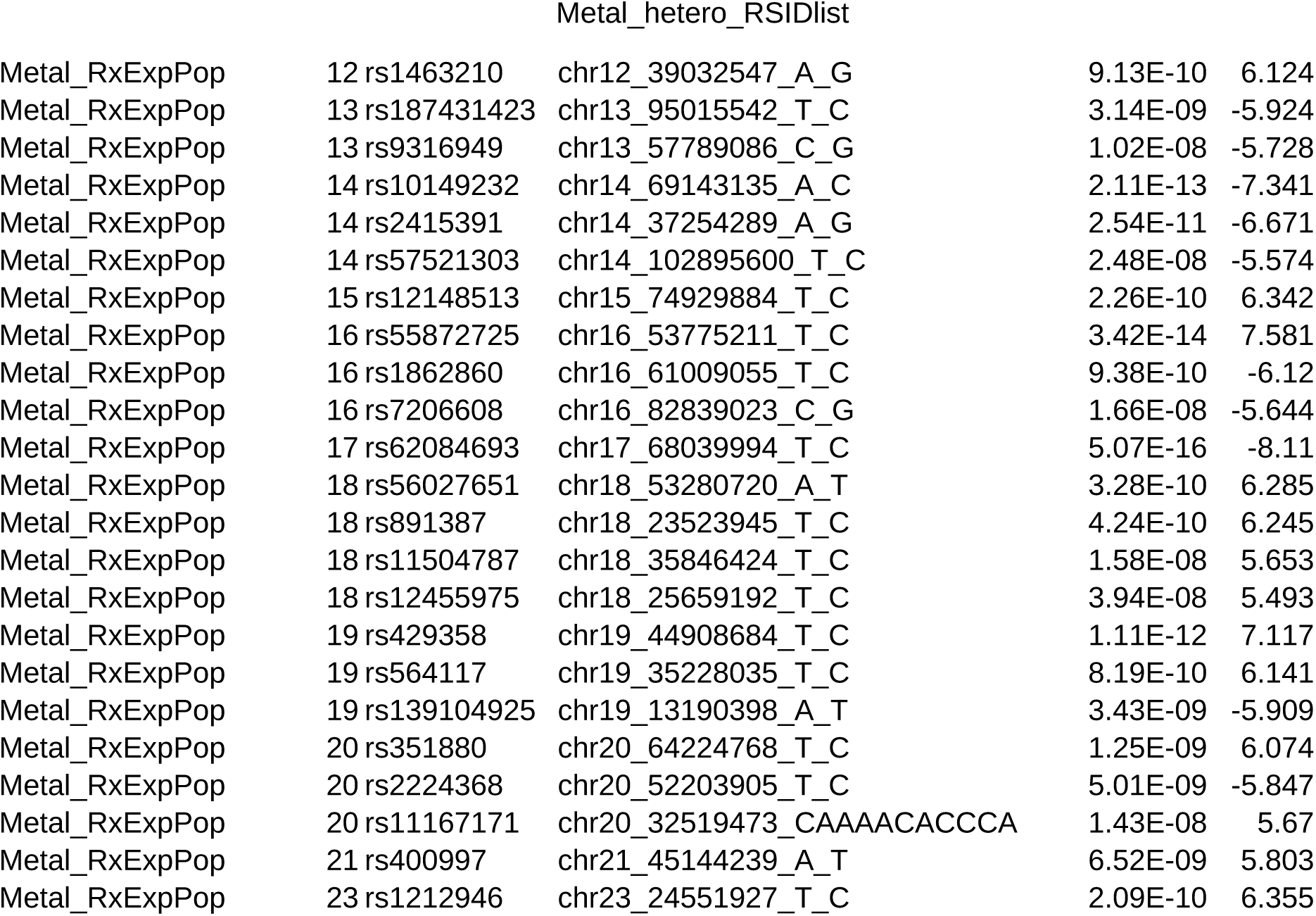
Lead SNPs of opioid prescription phenotypes with Open Targets annotation. The table lists the lead SNPs of each locus, defined by ±3 Mb regions, identified in the three base phenotypes of the meta-analysis: binary prescription status (RxExpPop), dosage plus binary prescription (RxDosePop), and dosage among users (RxDoseUsers). For each SNP, chromosome, region, rsID, and p value are provided. Open Targets annotation is included for each locus, showing the mapped gene, associated disease or trait, locus-to-gene (L2G) score, posterior probability, and study ID.

Locus overlap patterns showed closer relationships of RxExpPop with RxDosePop, while RxDoseUsers shared fewer loci with either trait (Fig. 3a). Meta-analysis GWAS closely mirrored biobank-level genetic architecture: across 27 comparisons between meta-analyses and single-biobank phenotypes, 18 showed high genetic correlation (rg > 0.90), with the lowest correlation observed between the RxExpPop meta-analysis and UKBB-RxDoseUsers (rg = 0.77; P = 7.22 × 10⁻⁴⁵; Fig. 3b; Supplementary Fig. 7).

**Figure 1.**
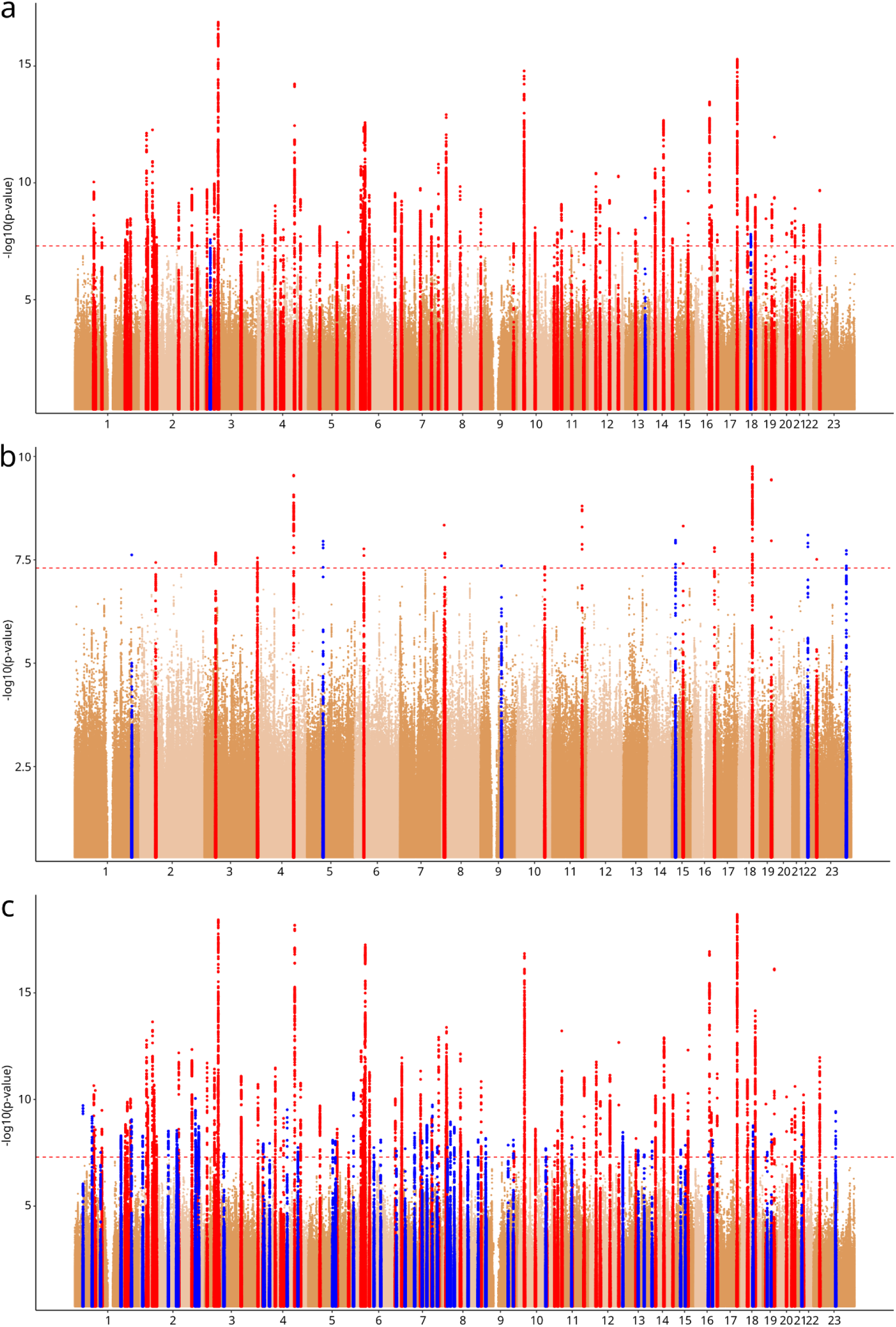
Genome-wide association results for three opioid prescription phenotypes. Manhattan plots of meta-analyses across biobanks for (a) binary opioid prescription status (RxExpPop; 78 independent lead SNPs), (b) dosage among users (RxDoseUser; 20 independent lead SNPs), and (c) combined opioid dosage plus binary prescription (RxDosePop; 135 independent lead SNPs). The red line denotes the genome-wide significance threshold (*P* < 5 × 10⁻⁸), Loci in blue are unique to the trait and loci marked in red are shared with at least one other trait.

**Figure 3.**
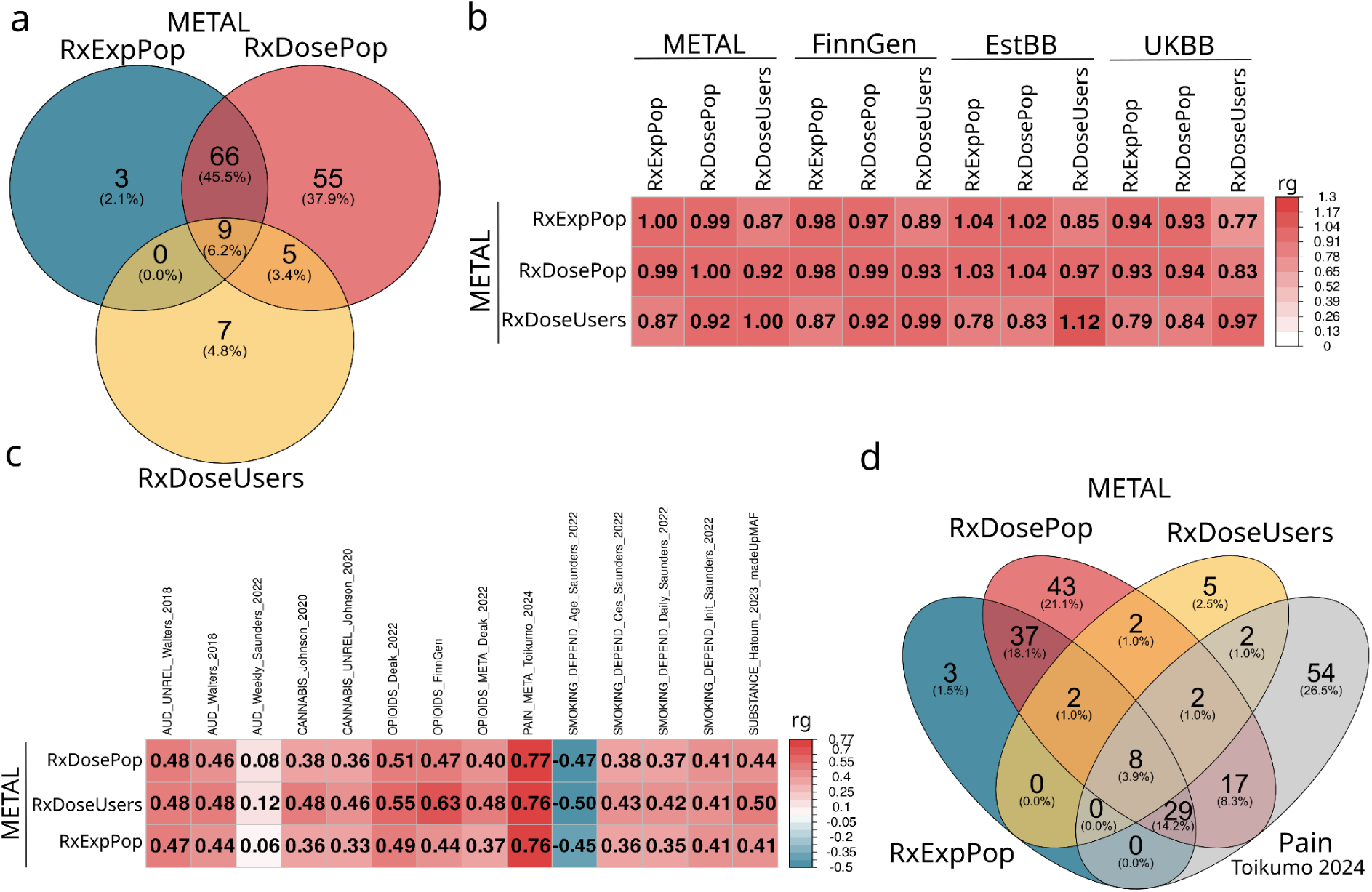
Overlap and genetic correlations of opioid prescription phenotypes. (a) Genetic correlations (rg) between METAL and individual biobank results (FinnGen, EstBB, UKBB). (b) SNP overlap among the three metal phenotypes: binary prescription status (RxExpPop), dosage plus binary prescription (RxDosePop), and dosage among users (RxDoseUser). (c) Genetic correlations of opioid phenotypes with published studies of alcohol use disorder^18,19^, cannabis use^21^, opioid dependence^8^, pain^17^, smoking behaviors^19^, and substance use^20^. (d) SNP overlap between opioid phenotypes and the recent pain GWAS (Toikumo et al. 2024)^17^.

Overall, these results highlight a robust, biologically coherent genetic architecture underpinning opioid prescribing, strongly replicable across multiple large-scale biobanks, and setting a foundation for further genetic and pathway analyses.

### Genetic risk loci implicate neuronal and metabolic pathways

We next annotated all lead variants using fine-mapping and functional consequence data from the Open Targets Genetics portal to identify their most likely target genes for all GWAS (Supplementary Table 8-11). We identified linked genes for 181 of 234 independent phenotype-loci signals (77.4%) and previously reported trait associations for 168 of 234 signals (71.8%; Supplementary Table 8). Of these, 65 (27.8%) overlapped with prior GWAS of pain or analgesic use, whereas others connected to weight (21 signals; 9.0%), substance-use behavior (10; 4.3%), socioeconomic factors such as education (7; 3.0%), and mental traits (6; 2.6%). Cardiometabolic traits (e.g., hypertension, diabetes) formed the largest remaining category (report exact n), underscoring pleiotropic links beyond pain pathways.

To prioritize the most likely causal genes, we combined fine-mapped credible sets with Locus-to-Gene (L2G) scores from Open Targets. Five genes had both an L2G score >0.9 and a posterior probability >0.5 in prior GWAS: ***CDH13*** and ***APOE*** (shared across all three phenotypes), ***COL11A1*** (shared by RxDosePop and RxExpPop), and ***LRP1B*** and ***ANAPC4*** (exclusive to RxDosePop). Three additional genes were found in all three phenotypes with high L2G scores: ***NCAM1*** (0.91), ***DCC*** (0.91), and ***MAML3*** (0.81). Other notable high-scoring candidates included ***PCDH17*** (0.96), ***KCNIP4*** (0.94), ***NEGR1*** (0.94), ***MKRN1*** (0.94), and ***PDE4B*** (0.93).

While the L2G score reflects how likely a variant is to regulate a gene across multiple lines of evidence, the posterior probability quantifies the likelihood that a specific variant is causal at its locus given fine-mapping results. Using posterior probability thresholds >0.9, we identified nine unique genes consistently across RxExpPop and RxDosePop: ***FTO*, *WSCD2*, *COX5A*, *ARHGAP15*, *ATP6V1G3*, *NOL4L*, *ALAS1*, *ANAPC4*** and ***PRAG1*** (Supplementary Fig. 12). ***PRAG1*** was the only gene supported by high posterior probability in both dosage-related phenotypes (RxDosePop and RxDoseUsers), suggesting a potential role specific to opioid dosage.

LD score regression indicated modest SNP-heritability across phenotypes: h²_SNP_ = 0.080 (SE = 0.003; 95% CI ≈ 0.074-0.086) for RxExpPop, 0.108 (0.0034; 95% CI ≈ 0.101-0.115) for RxDosePop, and 0.050 (0.0021; 95% CI ≈ 0.046–0.054) for RxDoseUsers (Supplementary Fig. 6, Supplementary Table 12). Partitioned heritability highlighted central nervous system (CNS) enrichment across all three phenotypes, with RxDosePop showing the strongest signal (P = 3.2×10⁻⁶), followed by RxExpPop (P = 3.8×10⁻⁴) and RxDoseUsers (P = 5.2×10⁻⁴), each significant after multiple-testing correction (Supplementary Fig. 7a, Supplementary Table 13). Within brain annotations, the cortex remained significantly enriched for RxDosePop (P = 1.8×10⁻³) whereas RxExpPop and RxDoseUsers did not retain significance after correction (Supplementary Fig. 7b). Several other regions showed nominal enrichment; notably, the cerebellum had the strongest (uncorrected) signal for RxDoseUsers (P = 0.016), suggesting a partly distinct genetic profile for dosage among users. Chromatin-based enrichment analyses converged on brain tissues: among the top 30 tissue-specific chromatin annotations, significant signals were predominantly brain-related (16 for RxDosePop, 9 for RxExpPop, 7 for RxDoseUsers; Supplementary Fig. 7c, Supplementary Table 13).

Together, these analyses show that meta-analyses identify hundreds of loci across opioid prescription phenotypes. Many of these variants overlap with pain and medication traits, while others link to metabolic and behavioral outcomes. Although overall SNP-heritability is modest, all phenotypes show enrichment in cortical brain tissues pointing to shared neuronal mechanisms alongside dosage-specific contributions.

### Opioid prescription genetics overlap strongly with pain and partially with addiction

To assess which diseases and medications have the strongest genetic relationship to opioid prescription, we estimated genetic correlations between each phenotype and 2,481 traits in FinnGen. After multiple-testing correction (q-values)^16^, we still detected a large number of significant associations: RxExpPop correlated with 1,152 traits, RxDosePop with 1,137 traits, and RxDoseUsers with 965 traits (Supplementary Table 14).

Because the top 20 correlations for RxExpPop all yielded q = 0, we selected the three phenotypes with the highest rg together with the FinnGen pain phenotype to facilitate comparison across all opioid traits. For RxExpPop, the strongest correlations were with low back pain (rg = 0.88, q = 5.0e-300), the aggregated pain phenotype (rg = 0.88, q = 0), codeine-tramadol prescription (rg = 0.95, q = 0), and paracetamol prescription (rg = 0.92, q = 0; Supplementary Fig. 8a). RxDosePop showed a very similar pattern: low back pain (rg = 0.88, q = 0), pain (rg = 0.86, q = 0), codeine-tramadol (rg = 0.97, q = 0), and paracetamol (rg = 0.89, q = 0; Supplementary Fig. 8b). RxDoseUsers also correlated with the same traits, although estimates were consistently lower: low back pain (rg = 0.82, q = 2.6e-282), pain (rg = 0.77, q = 0), codeine-tramadol (rg = 0.95, q = 0), and paracetamol (rg = 0.72, q = 1.4e-197; Supplementary Fig. 8c). Similar patterns were observed in all three cohorts.

While several mental health traits showed significant genetic correlations, most exhibited only modest overlap with opioid prescription (mean rg ≈ 0.6). The strongest mental health associations were observed for PTSD in RxDoseUsers (rg = 0.91, p = 6.9×10⁻¹²), stress-related disorders in RxDosePop (rg = 0.74, p = 1.6×10⁻⁴⁸), and stress-related disorders in RxExpPop (rg = 0.73, p = 4.9×10⁻⁴⁴). When ranked among all correlated traits, the first mental health phenotype appeared at position 7 for RxDoseUsers, 70 for RxDosePop, and 79 for RxExpPop, suggesting that the dose of opioid exposure is more genetically linked to mental health than the binary prescription status (FinnGen, Supplementary Fig. 8d-f; UKBB, Supplementary Fig. 9a-c; EstBB, Supplementary Fig. 9d-f).

We then compared opioid prescription phenotypes with external GWAS of pain and substance use. A recent large-scale pain meta-analysis^17^ was strongly correlated with all three opioid phenotypes: RxExpPop (rg = 0.76, p = 0), RxDoseUsers (rg = 0.76, p = 3.7e-280), and RxDosePop (rg = 0.77, p = 0, Supplement Table 13). By contrast, correlations with substance use phenotypes such as alcohol^18^ and drug use disorders^8,19–21^ were more modest, ranging from 0.3 to 0.5 across phenotypes (Fig. 3c; Supplementary Table 15). These patterns were replicated in FinnGen, EstBB and UKBB (Fig. 5b, Supplementary Fig. 10 a b).

At the locus level, overlap with the Toikumo pain meta-analysis^17^ was substantial. Using our definition of genome-wide significant loci (lead SNP with the lowest p-value ±3 Mb), 58 of 112 loci overlapped with our opioid prescription GWAS (Fig. 3d). In the European-only analysis, 49 of 83 loci overlapped (Supplementary Fig. 11a). We also observed strong concordance with loci previously associated with opioid addiction in Deak et al.^8^, with 11 of 18 loci replicated in our analyses (Supplementary Fig. 11b). Quantitatively, overlap with the pain meta-analysis was 41% for RxDosePop, 47% for RxExpPop, and 47% for RxDoseUsers.

Together, these analyses show that opioid prescription phenotypes share their strongest genetic overlap with pain traits, while only partially overlapping with addiction-related loci and substance use disorders. This indicates that the genetics of opioid prescription primarily reflect pain biology, with a secondary component linked to substance use.

### Machine-learning phenotypes identify overuse of opioid prescriptions

Because RxDoseUsers showed more distinct genetic correlations than RxExpPop and RxDosePop, we aimed to refine prescription phenotypes by developing machine-learning-based measures of overuse. Two complementary phenotypes were constructed: **RxOverUse_Onset**, capturing early onset of opioid prescription, and **RxOverUse_Amount**, reflecting excess dosage among individuals with comparable medical histories.

For RxOverUse_Onset, we trained XGBoost classifiers on the RxExpPop phenotype. Individuals were classified as early onset if exposed (RxExpPop = 1) but predicted as non-exposed (predicted = 0). The model showed stable performance, with mean squared errors (MSE) of 0.167-0.168, R² values of 0.317-0.322, and AUCs of 0.825-0.827 (Supplementary Figs. 13-15). Non-opioid pain medications were the strongest predictors, supporting that this phenotype primarily reflects pain-driven prescription.

For RxOverUse_Amount, XGBoost models were trained on the RxDoseUsers phenotype. Within opioid users, individuals were classified into overuse and underuse groups relative to their predicted dosage. Model performance was lower than for the binary phenotype, reflecting the greater complexity of the task, with MSE values of 3.07-3.08 and R² around 0.265. Overuse status was assigned conservatively when the predicted dosage exceeded the minimum observed opioid value in FinnGen (Supplementary Fig. 16).

We then conducted GWAS of both phenotypes. RxOverUse_Onset yielded no genome-wide significant associations (Fig. 4a). In contrast, RxOverUse_Amount identified a single locus on chromosome 22 (rs5751229, p=1.12e-8; Fig. 4b). This association was unique to RxOverUse_Amount within FinnGen, not appearing in the three base phenotypes, but it did replicate in the RxDoseUsers meta-analysis, confirming that the signal was genuine though not detectable in FinnGen alone.

**Figure 4.**
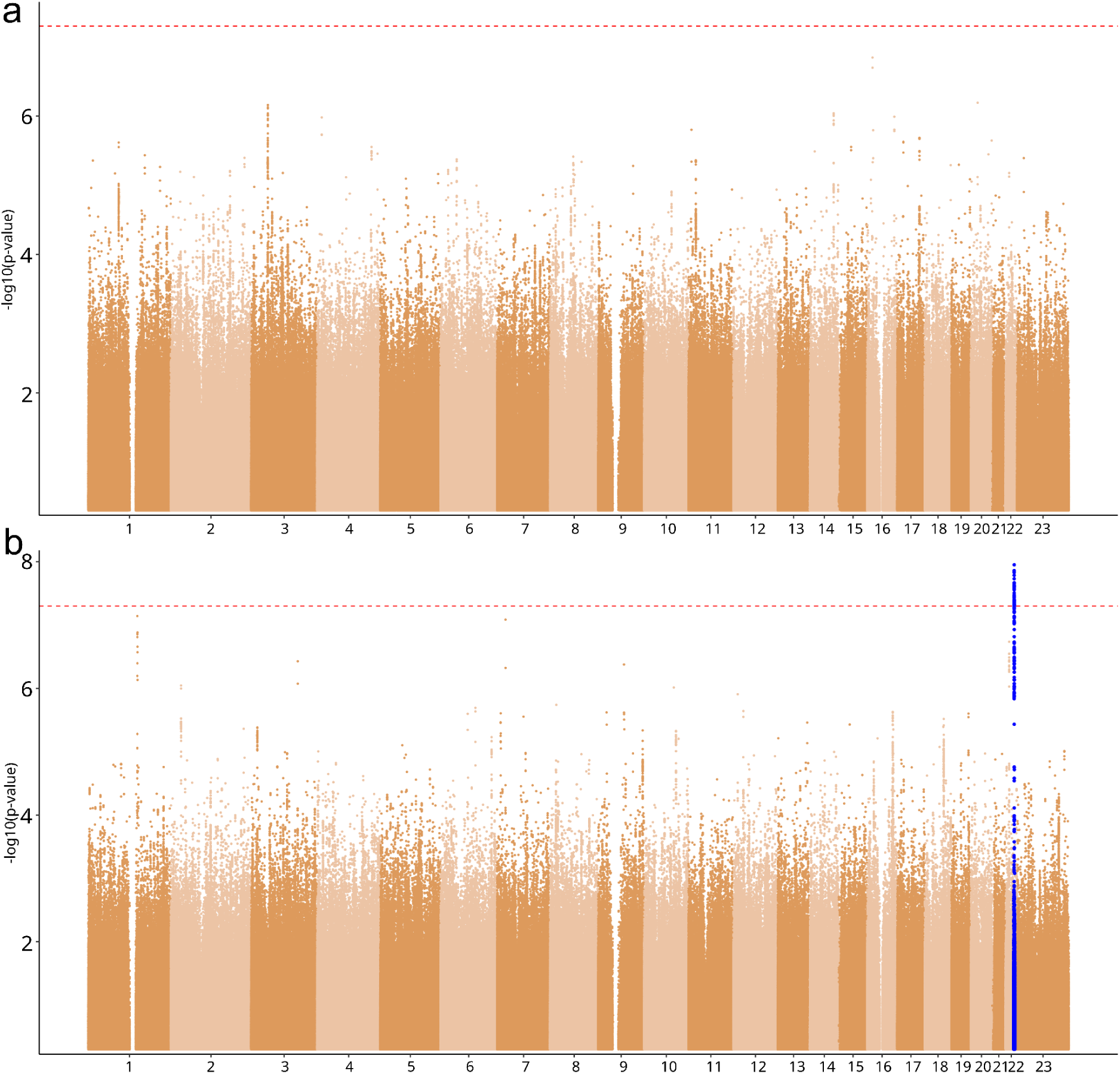
Genome-wide association results for machine learning–derived opioid overuse phenotypes. (a) *RxOverUse_Onset* phenotype, derived from misclassification in the binary opioid prescription model (RxExpPop). (b) *RxOverUse_Amount* phenotype, derived from predicted opioid intake based on the dosage model (RxDoseUsers). For the RxOverUse_Amount phenotype, we identified a genome-wide significant lead SNP on chromosome 22 (rs5751229). The red line denotes the genome-wide significance threshold (*P* < 5 × 10⁻⁸). Loci in blue are unique to the trait.

For the significant locus of RxOverUse_Amount, fine-mapping identified rs58099562 on chromosome 22. Annotation with Open Targets showed that this variant was part of multiple credible sets across published GWAS, including pain and insomnia, and it mapped primarily to *CYP2D6/CYP2D7*. This gene is central to drug metabolism and has established links to pain and substance use biology.

Together, these analyses demonstrate that machine learning can be used to construct valid overuse phenotypes from prescription data. While RxOverUse_Onset resembled the baseline exposure phenotype and did not identify novel loci, RxOverUse_Amount revealed a genome-wide significant association at rs5751229 that fine-mapped to rs58099562 at *CYP2D6/CYP2D7*. Importantly, this signal was unique to RxOverUse_Amount in FinnGen but was also observed in RxDoseUsers in the meta-analysis, highlighting its robustness and showing that ML-derived phenotypes can uncover dosage-specific genetic associations not evident in standard analyses.

### Overuse phenotypes separate pain-driven and addiction-driven genetic components

To understand what biology the machine-learning overuse phenotypes capture, we estimated genetic correlations of RxOverUse_Onset and RxOverUse_Amount with the three base opioid prescription phenotypes. RxOverUse_Onset correlated most strongly with population-wide measures (rg = 0.80 with RxExpPop; rg = 0.76 with RxDosePop) and more weakly with RxDoseUsers (rg = 0.59). In contrast, RxOverUse_Amount correlated most strongly with RxDoseUsers (rg = 0.77) and more weakly with RxExpPop (rg = 0.38) and RxDosePop (rg = 0.49) (Fig. 5a).

**Figure 5.**
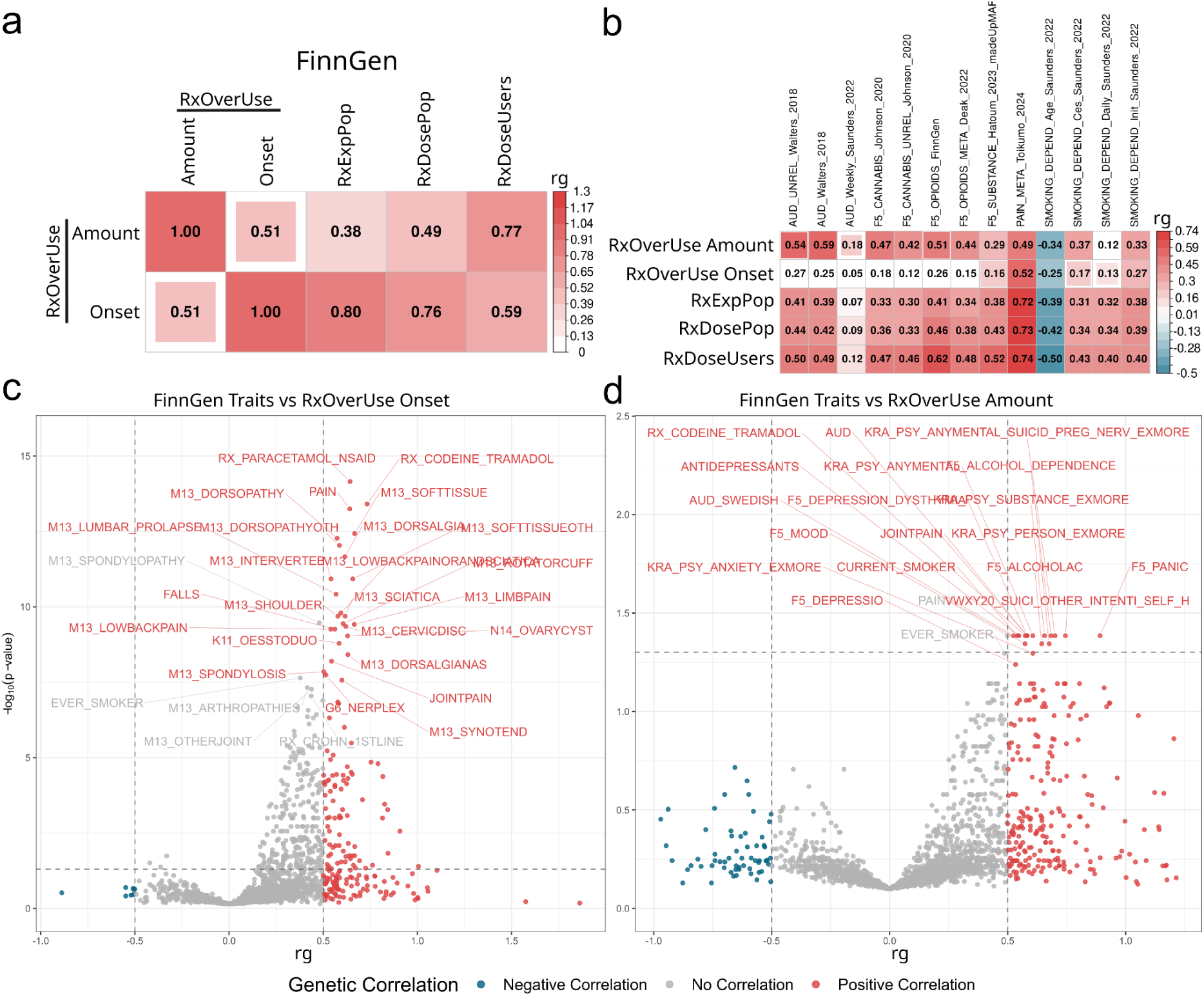
Genetic correlations of machine learning–derived opioid overuse phenotypes. (a) Genetic correlations (*rg*) between FinnGen overuse phenotypes (RxOverUse_Amount, RxOverUse_Onset) and prescription-based phenotypes (RxExpPop, RxDosePop, RxDoseUsers). (b) Genetic correlations of all FinnGen phenotypes with published studies of alcohol use disorder^18,19^, cannabis use^21^, opioid dependence^8^, pain^17^, smoking behaviors^19^, and substance use^20^. (c) Genetic correlations of FinnGen traits with RxOverUse_Onset, highlighting strong correlations with musculoskeletal pain, prescription pain medications, and related disorders. (d) Genetic correlations of FinnGen traits with RxOverUse_Amount, showing significant correlations with psychiatric, substance use, and pain-related traits.

To assess whether the overuse phenotypes capture genetic signals beyond those driving opioid prescription in general, we compared their genetic correlations with external GWAS of pain and substance use (Fig. 5b). RxOverUse_Onset showed broadly similar patterns to the main prescription phenotypes but with reduced power, with the strongest correlation observed for pain (rg = 0.52, p = 3.0e-8) and weaker correlations for other traits. By contrast, RxOverUse_Amount displayed a shifted profile: while still correlated with pain (rg = 0.49, p = 4.5e-5), it showed stronger correlations with substance use traits, including alcohol use disorder^18^ (rg = 0.59, p = 3.8e-3) and FinnGen opioid abuse Phenotype (rg = 0.51, p = 7.7e-3).

After observing that RxOverUse_Amount showed different correlation patterns than RxOverUse_Onset or the three base phenotypes in external reference studies, we asked whether this represented a broader shift in genetic architecture when examined across many traits. To address this, we estimated genetic correlations with all FinnGen Phenotypes, providing a dense set of clinical comparison points. RxOverUse_Onset showed its strongest correlations with pain-related traits: low back pain (rg = 0.54, q = 5.5e-10), pain (rg = 0.64, q = 5.7e-14), codeine-tramadol prescription (rg = 0.73, q = 3.9e-14), and paracetamol prescription (rg = 0.64, q = 7.0e-15) aligning with the pattern of the base Phenotypes (Fig. 5c). In contrast, RxOverUse_Amount was dominated by mental health and substance use traits, with pain-related traits ranking lower. For RxOverUse_Amount, the top genetic correlations were dominated by psychiatric and substance use traits (Fig. 5d). The strongest signals included panic disorder (rg = 0.89, q = 0.041; rank 1), suicidal behavior (rg = 0.75, q = 0.041; rank 2), suicide-related features (rg = 0.70, q = 0.041; rank 3), alcohol dependence (rg = 0.68, q = 0.041; rank 4), and substance use disorders (rg = 0.66, q = 0.041; rank 5). By contrast, pain-related traits were much less prominent, with only three appearing in the top 20: joint pain (rg = 0.58, q = 0.041; rank 9), codeine-tramadol prescription (rg = 0.57, q = 0.041; rank 10), and the aggregated pain phenotype (rg = 0.50, q = 0.041; rank 14).

In conclusion, RxOverUse_Onset resembles the baseline exposure phenotype and primarily reflects pain-driven opioid prescription, whereas RxOverUse_Amount shows stronger genetic overlap with psychiatric and substance use traits than with pain.

## Discussion

We conducted the first cross-biobank GWAS of prescription-derived opioid exposure and dose, harmonized to total cumulative oral morphine equivalents (OME), across approximately 860,000 participants. Meta-analysis identified 234 genome-wide significant signals mapping to 145 unique genomic loci. Exposure-oriented traits (RxExpPop, RxDosePop) shared most of their genetic architecture and overlapped prominently with known pain-related traits^17,22^, whereas cumulative dose among users (RxDoseUsers) captured partly distinct genetic determinants. Tissue and chromatin enrichments specifically implicated cortical neuronal circuits, while a model-adjusted “excess dose” phenotype highlighted pharmacokinetic biology at *CYP2D6*/*CYP2D7*. Collectively, these results establish a biologically coherent genetic architecture underlying medical opioid use, separating indication-driven exposure from dosage-specific liability.

The reproducibility of these genetic findings across three distinct healthcare systems underscores their robustness and portability. Genetic correlations between matched phenotypes were consistently high, with modest attenuation largely attributable to coding differences between ATC-based prescriptions in FinnGen^23^ and EstBB^24^ and BNF/text-based medication fields in UK-Biobank^25^. The broad reproducibility across distinct prescription-capture methods strongly indicates that these phenotypes capture true underlying biology rather than local healthcare system practices.

We observed that genetic liability to prescription opioid use closely mirrors the clinical patterns identified in our epidemiological analyses. Age and body mass index (BMI) contributed significantly to both opioid exposure and accumulated dose, reflecting increased chronic pain burden and multimorbidity^26^ with advancing age and higher BMI^9,27–29^. In contrast, sex effects were bidirectional: female sex was positively associated with the likelihood of receiving any opioid prescription but negatively associated with cumulative dose among users. This pattern aligns with prior evidence suggesting that gender influences pain perception, healthcare-seeking behavior, and opioid metabolism^30,31^. Additionally, we noted modest but consistent associations between opioid prescription and psychiatric disorders, reinforcing previously described genetic correlations between chronic pain and depression and underscoring the shared biological basis of pain, psychiatric disease, and multimorbidity^26,32^. These clinical correlations parallel our genetic findings, where the strongest genetic overlap was observed with pain-related traits, while dose-specific phenotypes showed partial convergence with psychiatric and substance-use liability.

At the gene level, prioritized genes identified through Open Target fine-mapping association and integrative annotations revealed biologically plausible mechanisms. Genes with strong support included *APOE* and *CDH13*, *APOE* with established associations to pain and neurological traits^17,33,34^, and *CDH13* implicated in GABA signaling^35^, suggesting potential involvement in pain regulation pathways^36^. Other high-confidence genes, such as *COL11A1*^17,33^ and *ANAPC4*^37^, have documented associations with pain phenotypes, while *DCC*^37–41^, and *MAML3*^22,42,43^ have also been linked to substance-use traits, underscoring shared neurobiological pathways between analgesic requirements and addiction vulnerability. Additionally, several genes were specifically connected to both pain and opioid abuse^8,44^, including *NCAM1*^38,44^ (across all three phenotypes), *FOXP2*^45^, *FTO*^46^ (RxExpPop and RxDosePop), and *PDE4B*^47^ (RxDosePop). Taken together, these findings strongly suggest that opioid prescription genetics reflect common biology underlying both analgesic need and behavioral vulnerabilities.

The polygenic architecture of opioid prescription extends beyond analgesic pathways. Many identified loci demonstrated pleiotropy, with associations extending to broader comorbidities such as obesity, cardiovascular diseases, and mental health conditions including substance abuse. These observations align with epidemiological evidence demonstrating tight interconnections among pain, psychiatric disorders, and multimorbidity^9,26,28,32^. Although overall SNP-based heritability estimates were modest, like other prescription GWAS^48^, partitioned enrichment analyses specifically highlighted cortical regions implicated in higher-order pain processing and opioid receptor action^49^. Cortical regions are central to pain processing^50^, undergo plastic changes in chronic pain^51^ and serve as primary sites of opioid receptor action^49^. They are also implicated in how emotions influence pain perception^52^. Thus, our results suggest that cortical neuronal circuits rather than peripheral nociceptive pathways likely shape prescribing behavior, reinforcing the complex interplay between pain modulation, emotional regulation, and opioid intake patterns.

Machine learning has become a standard tool for improving EHR-based phenotypes, with applications ranging from broadening case definitions^53^ and imputing diagnoses^54,55^ to predicting disease onset^56^, applying liability-scale models^57,58^, and incorporating family history^59,60^ or clinical records^61–63^. While these strategies highlight the potential of ML, they can also introduce noise^57^. Here, we applied ML in a different way: not to predict traits across cohorts, but to reclassify opioid intake within FinnGen relative to each individual’s comorbidity burden.

By applying machine-learning methods, we constructed two refined, model-adjusted phenotypes: early opioid initiation (RxOverUse_Onset) and excess opioid dose (RxOverUse_Amount). These phenotypes quantified deviations from expected opioid use patterns given each individual’s non-opioid medication and clinical history. One loci at rs5751229 reached significance in RxOverUse_Amount, which has also been associated in external GWAS of pain^17^, alcohol use disorder^19^, and smoking^19^, underscoring its potential role at the intersection of pain sensitivity and substance use. After finemapping the excess dose phenotype revealed a dosage-dependent genetic association at the *CYP2D6/CYP2D7* locus (rs58099562), strongly implicating pharmacokinetic variation. The locus appeared in multiple credible sets across published GWAS including pain^17^ and insomnia^64^. The variant mapped to *CYP2D6*/*CYP2D7*, with *CYP2D7* previously implicated in pain^17^ and *CYP2D6* linked to opioid pain^65,66^, opioid metabolism^67,68^, and opioid use disorder^67,69^, supporting a plausible biological link between opioid metabolism, dosage variation, and addiction risk. In the Finnish population, rs58099562 and rs3892097 are in very high linkage disequilibrium (r² ≈ 0.96, LDlink^70^).

The rs3892097 variant reduces CYP2D6 enzymatic activity and confers a poor-metabolizer phenotype for a wide range of drugs, including antidepressants^71^ and opioid analgesics. Codeine requires CYP2D6-mediated O-demethylation to be converted into its active form, morphine. Reduced CYP2D6 function therefore leads to lower morphine formation, as only a small fraction of codeine is normally activated through this pathway, resulting in diminished analgesic efficacy in poor metabolizers^15,66^. This mechanism provides a biologically coherent interpretation of the association between rs3892097 and higher opioid use.

Furthermore, the opioid dose showed stronger genetic correlations with mental health traits than with pain-related traits. Among the base phenotypes, mental health traits ranked more highly by genetic correlation for RxDoseUsers (top mental health trait rank = 7) than for RxDosePop (rank = 70) or RxExpPop (rank = 79). For the RxOverUse_Amount phenotype, nearly all top genetic correlations were with mental health traits, in contrast to the other opioid phenotypes, which primarily correlated with pain-related traits. This pattern suggests that while prescribed opioid use generally reflects underlying pain conditions, excessive opioid consumption may instead capture neurobiological factors related to pain perception and processing, such as pain tolerance, sensitivity, and vulnerability to addictive behaviors, linking opioid use more closely to mental health than to pain causation. While the underlying mechanisms remain unclear, these findings highlight a biologically meaningful connection between opioid dosage patterns and mental health that warrants further investigation.

While the structural variation at *CYP2D6* warrants targeted investigation beyond the scope of this study, these model-refined phenotypes captured dosage-specific genetic liability distinct from traditional indication-driven definitions. Together, they underscore the potential of integrative computational approaches to refine EHR-derived traits and reveal new biological dimensions of opioid use.

This study has several methodological strengths. By integrating data from three independent biobanks operating within distinct healthcare systems, we achieved robust replication and broad generalizability of the findings. The harmonization of opioid prescriptions into oral morphine equivalents (OME) allowed consistent meta-analysis across diverse drug formulations, while the combined use of traditional and model-adjusted phenotypes disentangled genetic components related to indication versus dosage. Together with extensive replication and stringent heterogeneity filters, these design choices minimized false positives and strengthened the biological validity of our results.

Nonetheless, several limitations should be acknowledged. Our analyses were restricted to individuals of European ancestry, underscoring the need for future replication in more diverse populations to assess portability. Prescription records capture dispensing rather than actual consumption and do not account for illicit opioid use or adherence variability. Residual confounding by indication, particularly differences in pain severity, may persist despite partial adjustment in the model-based phenotypes. Finally, cross-biobank variation in drug coding systems (ATC vs. BNF) could introduce minor inconsistencies, although high genetic correlations between cohorts suggest this effect is limited.

## Conclusion

Recent work has shown that genetic analyses of medication-prescription data can provide meaningful insights into underlying biology, and our results further support this idea^72^. Our analyses demonstrate that prescription-derived opioid phenotypes contain multiple layers of genetic signal that can be disentangled with careful modeling. By harmonizing opioid prescribing data across almost 860,000 individuals and integrating machine-learning–based phenotype refinement with genome-wide analyses, we show that standard prescription measures primarily capture pain-related biology, whereas disproportionately high dosing reflects a distinct genetic liability enriched for psychiatric, substance-use, and pharmacokinetic pathways. Notably, the excess-dose phenotype revealed a clear association at the CYP2D6/CYP2D7 locus, consistent with known variation in opioid metabolism, highlighting how data-driven machine learning based phenotyping can bring dosage-specific mechanisms into focus.

More broadly, this work illustrates the value of combining large-scale real-world prescribing data with machine-learning approaches to capture clinically meaningful variation not evident in traditional traits. Such strategies may improve the detection of genetic influences on medically prescribed substances and help identify individuals with atypical opioid requirements, informing safer and more individualized pain-management practices.

## Methods

### Cohort Information: FinnGen, Estonian Biobank, and UK Biobank

This study utilizes data from three large-scale biobanks, FinnGen, the Estonian Biobank (EstBB), and the UK Biobank (UKBB). Each combines genotype data with longitudinal health records.

FinnGen^23^ Data Freeze 12 (DF12) includes 520,000 participants, about 10% of the Finnish population. The cohort is enriched for disease cases, with mean age 53 years and 43% men. National registries provide hospital, outpatient, primary care, cancer, prescription, reimbursement, and demographic data. Additional information includes smoking, occupation, and number of children. In this study, 497,603 individuals were analyzed, of whom 281,529 had at least one opioid prescription.

The EstBB^24^ has 200,000 participants, ∼20% of the Estonian adult population. Recruitment reflects national age, sex, and regional structure. Registry linkage gives longitudinal data on diagnoses, treatments, and prescriptions. We analyzed 208,376 individuals, including 65,922 with at least one opioid prescription.

The UKBB^25^ includes ∼500,000 participants with health data from hospital and primary care records. These cover diagnoses, procedures, medications, and lifestyle. Medication data are coded with the British National Formulary (BNF) system, in contrast to ATC coding in FinnGen and EstBB. In UKBB, we included only 184,597, who had at least one BNF-coded prescription in the records (n=184,597). The GWASs were performed in the ‘White British’ self-reported ethnic background subset (Data-Field 21000), totaling 153,696d, of whom 54,313 had at least one opioid prescription in the records.

All three biobanks provide genotypes for 700,000-800,000 markers per individual. Prescription data were available in all cohorts, allowing pharmacogenetic analysis across healthcare systems. Variant and sample quality control procedures are described in the Supplementary Methods.

### Population Characteristics

We evaluated demographic and clinical factors associated with opioid prescription and cumulative dose in FinnGen using regression-based analyses. Two complementary models were applied: a binary model for RxExpPop and a continuous model for RxDoseUsers.

For RxExpPop, we used logistic regression with opioid intake as the dependent variable and each covariate as an independent variable in separate models. All models were adjusted for age and sex. Odds ratios (ORs) and corresponding 95% confidence intervals (CIs) were derived by exponentiating the regression coefficients (OR = exp(β)), providing an interpretable measure of the change in odds of opioid use per unit increase in each predictor.

For RxDoseUsers, the total oral morphine equivalent (OME) was log_n_-transformed to approximate normality, and linear regression was applied with log(OME) as the dependent variable. Each comorbidity or demographic variable was modeled separately, with age and sex included as covariates. Regression coefficients (β) reflect the expected change in log-transformed cumulative opioid dose per unit increase in the predictor, conditional on age and sex.

All analyses were performed in R (v4.3.2). P-values < 0.05 were considered nominally significant, and 95% CIs were reported where applicable to convey estimate precision.

### Opioid Conversion Factors

In FinnGen, opioid exposure was constructed from pharmacy-dispensation records identified by the national product code (VNR) using catalogues from the Finnish Medicines Agency (Fimea)^73^. Product strength and package size were first harmonized: composite entries (for example “5 × 10 ml”) were converted into a single numeric amount (“50 ml”), salt forms and fixed-dose combinations were standardized, and duplicate or inconsistent entries were resolved into unified fields. When strength or package size information was missing after harmonization, records were completed for rare outliers using other national formularies^74–76^. For rare or outdated products that could not be located, the nearest common presentation of the same molecule and strength range was substituted, using conservative estimates to avoid inflating the dose.

For each entry, the total dose of opioid base was calculated by multiplying normalized strength in milligrams with the total purchased (FinnGen and EstBB) or prescribed (UKBB) quantity. Transdermal formulations recorded as release rates were converted into milligrams per patch based on labelled wear time (72 hours for fentanyl, 168 hours for buprenorphine, or 96 hours when specified). This ensured that oral, parenteral, and transdermal routes could be expressed on the same milligram scale.

Each molecule and route of administration was then assigned a conversion factor reflecting potency relative to morphine. Factors were primarily taken from published comparative guidelines (Nielsen et al., 2014)^77^. When route-specific values were not available, equivalent routes were treated together (for example, buccal, sublingual, lozenge/oromucosal, and nasal). In cases where no published factor existed but pharmacology supported a simple relation, a conservative assumption was applied. For example, parenteral tramadol was set to be approximately twice as strong as oral tramadol, consistent with clinical conversion charts (Gloucestershire Hospitals NHS Foundation Trust)^78^.

### Opioid Values

The opioid value phenotype was constructed from longitudinal prescription data in FinnGen (DF12), EstBB, and UKBB. In FinnGen and EstBB, prescription records were extracted from the national drug purchase registries, restricted to medications with ATC codes beginning with *N02A* (opioids). In EstBB, the code *N07BC02* was additionally included to capture methadone prescriptions, which had previously been listed under *N02AC02*. Codes *N02AB02* (pethidine), *N02AB01* (ketobemidone) and *N02AX06* (tapentadol) were excluded in EstBB due to insufficient prescription numbers and lacking opioid information.

To document which medications were included in each study, we generated summary tables showing the number of prescriptions and exposed individuals per drug (Supplementary Tables 4-6). In FinnGen, to maintain anonymisation, all counts of ≤5 events or individuals were reported as 6.

For each individual (i), the opioid value (OV) was calculated as the sum of prescription-level exposures:

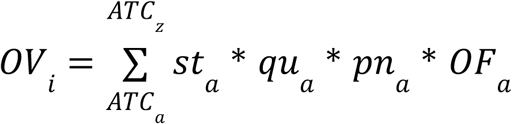

where *st* is opioid strength, *qu* is package quantity, *pn* is the number of prescriptions, and *OF* is the opioid factor. This definition combines drug potency with prescription volume, providing a continuous measure of exposure.

In both UKBB and EstBB, the same harmonisation strategy was applied to ensure comparability with FinnGen. Where published opioid factors were missing or uncertain, conservative rules were used: drugs likely to be stronger than morphine were capped at morphine equivalence, and uncertain weak or partial agonists were treated as equivalent to codeine. These rules avoided overestimation and ensured consistent calibration across cohorts.

### Opioid Phenotypes

After calculating opioid values for each individual in each biobank, three phenotypes were derived. Because opioid values were highly skewed, all continuous traits were log_n_-transformed to improve normality and ensure suitability for genome-wide association studies (GWAS).

- **RxExpPop**: a binary phenotype indicating whether an individual had ever been prescribed an opioid (1 = yes, 0 = no).
- **RxDoseUsers**: a continuous phenotype of total opioid dose among individuals with at least one prescription.
- **RxDosePop**: a continuous phenotype of total opioid dose across the entire population, assigning a value of zero to individuals without prescriptions. This hybrid phenotype combines exposed and unexposed individuals, resulting in a Tweedie-like distribution.

Together, these phenotypes capture complementary aspects of opioid use: initial exposure, prescribed dose among users, and dose distribution in the population as a whole.

### Genome-wide Association Study (GWAS)

In FinnGen, GWAS was performed using Regenie v2.2.4 for both binary and quantitative phenotypes. In EstBB, Regenie v3.2 was applied and UKBB Regenie v4.1^79^. Covariates in all cohorts included sex, age at death or end of follow-up, age², the first ten genetic principal components, and biobank-specific genotyping batch and chip information. Variants were filtered to those with minor allele frequency (MAF) > 0.001 and imputation info score > 0.8.

The three opioid phenotypes were analyzed within this framework. RxExpPop, representing any opioid prescription versus none, was modeled with logistic regression (Supplementary Fig. 17a). RxDoseUsers, reflecting cumulative opioid dose among exposed individuals, was log_n_-transformed and modeled with linear regression, which approximated normality (Supplementary Fig. 17b). RxDosePop, representing dose across the full population, exhibited a Tweedie-like distribution due to the mixture of non-users and skewed positive values (Supplementary Fig. 17c). Although linear regression is generally robust to skewness in large samples^80,81^, the recommended approach is inverse-rank normalization. To assess robustness, GWAS results from the Tweedie model were compared to those obtained after inverse-rank transformation and found to be nearly identical (genetic correlation rg = 0.9993, P = 0). In UK Biobank, opioid dose values were originally log_10_-transformed, whereas FinnGen and Estonian Biobank used natural log transformation. This discrepancy affects only the scale of effect size estimates (β and SE), not the underlying Z-scores or P-values. Accordingly, it had no impact on downstream analyses, including genetic correlation and meta-analysis, as both rely on standardized test statistics. To confirm this, UK Biobank phenotypes were re-transformed to the natural logarithm scale (multiplied by ln(10)) and reanalyzed; results were numerically indistinguishable from the original log_10_-based GWAS. On this basis, the log-transformed phenotype model was retained, as it preserved interpretability of opioid dose values while yielding equivalent results across cohorts.

Meta-analysis was performed across the three biobanks using METAL^82^ (release 2020-05-05). Fixed-effects inverse-variance weighted models were applied, and heterogeneity was assessed with Cochran’s Q statistic and the I² metric. Variants with heterogeneity p < 0.05 were excluded from downstream analyses.

### GWAS preparation of 3rd party studies

To contextualize our findings within existing literature, we integrated and harmonized external genome-wide association studies (GWAS) for comparison. Given that GWAS results are published in different genomic builds, we performed a liftover conversion from hg19 to GRCh38 for all datasets originally mapped to hg19. This was achieved using a local installation of the UCSC Liftover tool^83^. For SNPs missing reference SNP cluster IDs (rsIDs), we imputed rsIDs where possible to ensure consistency across datasets.

### Fine-mapping

To identify credible sets of variants underlying Finnish GWAS signals, we applied Sum of Single Effects (SuSiE) algorithm (v.0.11.92)^84^ using the FinnGen LD reference panel (fin_ldsc). Regions were defined as 3 Mb windows around genome-wide significant lead variants (P < 5 × 10⁻⁸), with overlapping windows merged; the MHC was excluded due to its complexity. In-sample linkage disequilibrium was computed from dosages using LDstore2^85^. Fine-mapping was conducted with SuSiE, allowing up to ten causal variants per locus (L = 10). The posterior inclusion probabilities (PIP) were reported for each variant, from which 95% and 99% credible sets were derived.

### SNP-based heritability and genetic correlation

SNP-based heritability and cross-trait genetic correlations were estimated using LD Score Regression (LDSC v1.0.1)^86^ in FinnGen using the FinnGen “fin_ldsc” reference panel, which captures Finnish-specific LD structure. For European ancestry datasets, precalculated LD scores were derived from the 1000 Genomes European reference^87^. Summary statistics were preprocessed with restricting to HapMap3^88^ SNPs.

For FinnGen-wide phenotype correlations, multiple testing was controlled using Storey’s q-value procedure^16^ (qvalue, version 2.4.2), which estimates the false discovery rate (FDR). This approach is less conservative than Bonferroni correction and is appropriate given the high correlation between many phenotypes (e.g. disease categories and subtypes).

Partitioned SNP heritability was estimated using stratified LDSC (S-LDSC)^89,90^, with summary statistics preprocessed to include only HapMap3 SNPs^88^. Three complementary models were applied: a cell baseline model to assess enrichment across tissue categories^89^, a brain baseline model to identify specific brain regions enriched for genetic signal^90,91^, and a tissue-specific model incorporating chromatin and genetic data^90^ For all reference baseline models, multiple testing was controlled using Bonferroni correction to ensure rigorous error control across annotations. For plotting vulcanoplots we set the manual floor p(0)=min(p>0).

### Lead SNP and gene annotation

Lead SNPs from the meta-analysis GWAS were defined as the variants with the lowest p-value within each locus, with loci assigned as ±1.5 Mb around the lead SNP (3 Mb window). Lead variants were annotated using fine-mapping and functional consequence data from the Open Targets Genetics Portal (https://genetics.opentargets.org, API v4)^92^. For each variant, fine-mapping information was retrieved through the Open Targets GraphQL API, including the study identifier, trait description, effect size (β), *p*-value, posterior probability, and locus-to-gene (L2G) score^93^. When multiple associations were available for a variant, the fine-mapped signal with the highest posterior probability was retained. If several studies contained the same variant, preference was given to those reporting pain-related traits; otherwise, the association with the strongest statistical evidence (lowest *p*-value) was selected. The resulting annotations provided trait-level, study-level, and gene-level information, including Ensembl gene identifiers and HGNC-approved symbols (Table 2).

Variants for which no fine-mapping data were available were annotated using the Ensembl Variant Effect Predictor (VEP) via the Open Targets API^92^. Among all predicted variant-gene consequences, the annotation with the highest predicted impact severity (HIGH > MODERATE > LOW > MODIFIER) and the smallest distance from the transcription start site was selected. This provided a functional consequence-based fallback annotation for variants outside credible sets.

All results were combined into comprehensive variant-level annotation tables and exported as tab-delimited files. This approach ensured consistent integration of fine-mapping and functional consequence data, enabling systematic gene prioritization for all genome-wide significant variants (Supplementary Table 14).

### Data Preparation

For the prediction analysis, we trained a model on opioid values extracted from 6 opioid medication codes (N02AX, N02AJ, N02AA, N02AE, N02AC, N02AB) across 520,149 Finnish individuals.

For the prediction we used all available information in FinnGen, and excluded opioid values for six opioid medication classes. A total of 9,617 phenotypes and covariates were included as predictors, covering disease phenotypes, prescription records, covariates, and socioeconomic factors. Most predictors were phenotype data, including the age of onset for each condition (n=4,006). Additionally, 516 different prescribed medications were included. Other predictors consisted of genetic principal components (PCs 1-10), demographic factors (age, sex, region of birth), education (degree, field of study), profession codes, and general lifestyle information such as weight, height, smoking status, and number of children.

For the prediction models, only the six opioid medication classes were excluded to provide a detailed view of individuals’ prescription patterns and phenotype history.

### Opioid Prediction and Overuse Phenotypes

Predictive models were trained using the XGBoost algorithm (XGBRegressor in Python)^94^, with *reg:squarederror* for continuous traits and *binary:logistic* for binary traits. Hyperparameters were optimized with RandomizedSearchCV in scikit-learn, using 5% of the training data for tuning. Models were trained with an 80/20 train-test split and evaluated across six independent runs (three for opioid dosage and three for opioid exposure). Performance was assessed using mean squared error (MSE), R², and area under the ROC curve (AUC). For continuous models, MSE ranged from 3.07 to 3.08 with R² values of 0.26-0.27. For binary models, MSE ranged from 0.16 to 0.17 with R² of 0.317-0.322 and AUC values of 82.3-82.6%. Feature importance was quantified using SHAP values (Supplementary Figs. 13–16).

Two opioid overuse phenotypes were derived from these models:

**RxOverUse_Amount**: This phenotype was based on predicted opioid intake among users (RxDoseUsers). Three separate dosage models were trained, and predictions were averaged to reduce variance. Individuals with an average predicted intake above the minimum observed opioid dose (log_n_(39)) were classified as overusers. The minimum observed opioid dose was defined as log n(39), corresponding to the smallest OME value assigned in FinnGen; this ensured that individuals classified as overusers exceeded at least one unit of OME above the minimum observed dose. The analysis was restricted to individuals with at least one recorded opioid prescription. Controls were defined as users with predicted intake at or below the threshold, excluding individuals with prediction variance >0.25. This yielded 3,163 over-users and 277,176 controls.
**RxOverUse_Onset**: This phenotype was based on exposure misclassification from binary models of opioid use (RxExpPop). Three models were trained, and predictions were averaged. The optimal threshold was determined using the Youden Index from the ROC curve. Overusers were defined as individuals with at least one opioid prescription (true RxExpPop = 1) who were incorrectly predicted as non-users. Controls consisted of individuals with no opioid prescriptions (true RxExpPop = 0), excluding those predicted as users (“underusers”). This yielded 67,307 over-users and 175,397 controls.

### Ethics

Study subjects in FinnGen provided informed consent for biobank research, based on the Finnish Biobank Act. Alternatively, separate research cohorts, collected prior the Finnish Biobank Act came into effect (in September 2013) and start of FinnGen (August 2017), were collected based on study-specific consents and later transferred to the Finnish biobanks after approval by Fimea (Finnish Medicines Agency), the National Supervisory Authority for Welfare and Health. Recruitment protocols followed the biobank protocols approved by Fimea. The Coordinating Ethics Committee of the Hospital District of Helsinki and Uusimaa (HUS) statement number for the FinnGen study is Nr HUS/990/2017.

The FinnGen study is approved by Finnish Institute for Health and Welfare (permit numbers: THL/2031/6.02.00/2017, THL/1101/5.05.00/2017, THL/341/6.02.00/2018, THL/2222/6.02.00/2018, THL/283/6.02.00/2019, THL/1721/5.05.00/2019 and THL/1524/5.05.00/2020), Digital and population data service agency (permit numbers: VRK43431/2017-3, VRK/6909/2018-3, VRK/4415/2019-3), the Social Insurance Institution (permit numbers: KELA 58/522/2017, KELA 131/522/2018, KELA 70/522/2019, KELA 98/522/2019, KELA 134/522/2019, KELA 138/522/2019, KELA 2/522/2020, KELA 16/522/2020), Findata permit numbers THL/2364/14.02/2020, THL/4055/14.06.00/2020, THL/3433/14.06.00/2020, THL/4432/14.06/2020, THL/5189/14.06/2020, THL/5894/14.06.00/2020, THL/6619/14.06.00/2020, THL/209/14.06.00/2021, THL/688/14.06.00/2021, THL/1284/14.06.00/2021, THL/1965/14.06.00/2021, THL/5546/14.02.00/2020, THL/2658/14.06.00/2021, THL/4235/14.06.00/2021, THL/4990/14.02.00/2023 Statistics Finland (permit numbers: TK-53-1041-17 and TK/143/07.03.00/2020 (earlier TK-53-90-20) TK/1735/07.03.00/2021, TK/3112/07.03.00/2021) and Finnish Registry for Kidney Diseases permission/extract from the meeting minutes on 4th July 2019.

The Biobank Access Decisions for FinnGen samples and data utilized in FinnGen Data Freeze 13 include: THL Biobank BB2017_55, BB2017_111, BB2018_19, BB_2018_34, BB_2018_67, BB2018_71, BB2019_7, BB2019_8, BB2019_26, BB2020_1, BB2021_65, BB22-0025-A01, BB22-0025-A03, BB23-0222-A01, BB22-0025-A04, BB22-0025-A06, BB22-0025-A08, THLBB2024_30. Finnish Red Cross Blood Service Biobank 7.12.2017, 13.11.2023, 001-2023, Helsinki Biobank HUS/359/2017, HUS/248/2020, HUS/430/2021 §28, §29, HUS/150/2022 §12, §13, §14, §15, §16, §17, §18, §23, §58, §59, HUS/128/2023 §18, BB22-0025-A01, BB22-0025-A02, BB22-0025-A05, BB22-0025-A07, BB22-0025-A09, BB22-0025-A10, BB22-0025-A03, BB23-0222-A01, BB22-0025-A04, BB22-0025-A06, BB22-0025-A08, Amendment_BB22-0025-A05, Decision allowing to continue data processing until 31st Aug 2027: BB_2021-0140, HUS/150/2022 §12, BB_2021-0139, HUS/150/2022 §13, BB_2021-0161,HUS/150/2022 §14, BB_2021-0164, HUS/150/2022 §15, BB_2021-0169, HUS/150/2022 §16, BB_2021-0170, HUS/150/2022 §17, BB_2021-0179, HUS/150/2022 §18, BB_2022-0262, HUS/150/2022 §58, BB22-0067, HUS/150/2022 §59, Auria Biobank AB17-5154 and amendment #1 (August 17 2020) and amendments BB_2021-0140, BB_2021-0156 (August 26 2021, Feb 2 2022), BB_2021-0169, BB_2021-0179, BB_2021-0161, AB20-5926 and amendment #1 (April 23 2020) and it’s modifications (Sep 22 2021), BB_2022-0262, BB_2022-0256, BB22-0025-A01, BB22-0025-A02, BB22-0025-A03, BB23-0222_A01, BB22-0025-A02, BB22-0025-A05, BB22-0025-A07, BB22-0025-A09, BB22-0025-A10, BB22-0025-A03, BB23-0222-A01, BB22-0025-A04, BB22-0025-A06, BB22-0025-A08, Decision allowing to continue data processing until 31st Aug 2027: AB20-5926, BB_2021-0140, BB_2021-0156, BB_2021-0161, BB_2021-0161, BB_2021-0164, BB_2021-0169, BB_2021-0179, BB_2022-0262, Biobank Borealis of Northern Finland_2017_1013, 2021_5010, 2021_5010 Amendment, 2021_5018, 2021_5018 Amendment, 2021_5015, 2021_5015 Amendment, 2021_5015 Amendment_2, 2021_5023, 2021_5023 Amendment, 2021_5023 Amendment_2, 2021_5017, 2021_5017 Amendment, 2022_6001, 2022_6001 Amendment, 2022_6006 Amendment, 2022_6006 Amendment_2, BB22-0067, 2022_0262, 2022_0262 Amendment, BB22-0025-A01, BB22-0025-A02, BB22-0025-A05, BB22-0025-A07, BB22-0025-A09, BB22-0025-A10, BB22-0025-A03, BB23-0222-A01, BB22-0025-A04, BB22-0025-A06, BB22-0025-A08, Decision allowing to continue data processing until 31st Aug 2027: BB/2021/5015, BB/2021/5017, BB/2021/5018, BB/2021/5023, BB/2022/6006, BB/2022/6001, BB/2022-0262, BB/2021/5010, Biobank of Eastern Finland 1186/2018 and amendment 22§/2020, 53§/2021, 13§/2022, 14§/2022, 15§/2022, 27§/2022, 28§/2022, 29§/2022, 33§/2022, 35§/2022, 36§/2022, 37§/2022, 39§/2022, 7§/2023, 32§/2023, 33§/2023, 34§/2023, 35§/2023, 36§/2023, 37§/2023, 38§/2023, 39§/2023, 40§/2023, 41§/2023, BB22-0025-A01, BB22-0025-A02, BB22-0025-A05, BB22-0025-A07, BB22-0025-A09, BB22-0025-A10, BB22-0025-A03, BB23-0222-A01, BB22-0025-A04, BB22-0025-A06, BB22-0025-A08, Decision allowing to continue data processing until 31st Aug 2027: MO-BB_2021-0179-A0, MO-BB_2021-0156_PRE-A01, BB_2021-0140, MO-BB_2021-0170_PRE-A0, MO-BB_2021-0169-A01, MO-BB_2022-0256-A01, MO-BB_2021-0161-A01, MO-BB_2021-0161-A02, BB22-0067-A01, MO-BB_2022-0262-A0, Finnish Clinical Biobank Tampere MH0004 and amendments (21.02.2020 & 06.10.2020), BB2021-0140 8§/2021, 9§/2021, §9/2022, §10/2022, §12/2022, 13§/2022, §20/2022, §21/2022, §22/2022, §23/2022, 28§/2022, 29§/2022, 30§/2022, 31§/2022, 32§/2022, 38§/2022, 40§/2022, 42§/2022, 1§/2023, BB2021-0140, BB22-0025-A01, BB_2021-0161, BB22-0025-A02, BB22-0025-A05, BB22-0025-A07, BB22-0025-A09, BB22-0025-A10, BB22-0025-A03, BB23-0222-A01, BB22-0025-A04, BB22-0025-A06, BB22-0025-A08, Decision allowing to continue data processing until 31st Aug 2027: BB_2021-0140, BB_ 2021-0161, BB_ 2021-0179, BB_ 2021-0156, BB_ 2021-0169, BB_ 2021-0170, BB22-0067-A01, Central Finland Biobank 1-2017, BB_2021-0169, BB_2021-0179, BB_2022-0256, BB_2022-0262, Decision allowing to continue data processing until 31st Aug 2027 for projects: BB_2021-0179, BB22-0067,BB_2022-0262, BB_2021-0170, BB_2021-0164, BB_2021-0161, and BB_2021-0169, BB22-0025-A01, BB22-0025-A02, BB22-0025-A05, BB22-0025-A07, BB22-0025-A09, BB22-0025-A10, BB22-0025-A03, BB23-0222-A01, BB22-0025-A04, BB22-0025-A06, BB22-0025-A08, Terveystalo Biobank STB 2018001 and amendment 25th Aug 2020, Finnish Hematological Registry and Clinical Biobank decision 18th June 2021, Amendment 2nd January 2024 and Arctic biobank P0844: ARC_2021_1001, ARC_2023_3003 (BB22-0025-A01), BB22-0025-A03, BB23-0222-A01, BB22-0025-A04, BB22-0025-A06, BB22-0025-A08.

The activities of the EstBB are regulated by the Human Genes Research Act, which was adopted in 2000 specifically for the operations of the EstBB. Individual level data analysis in the EstBB was carried out under ethical approval “1.1-12/624” from the Estonian Committee on Bioethics and Human Research (Estonian Ministry of Social Affairs), using data according to release application “6-7/GI/28554” from the Estonian Biobank.

Ethics approval for the UK Biobank study was obtained from the North West Centre for Research Ethics Committee (11/NW/0382). UK Biobank data used in this study were obtained under application 22627.

## Data Availability

All non individual data produced in the present study are available upon reasonable request to the authors

## Acknowledgement

The author gratefully acknowledges funding support from the University of Helsinki Research Foundation and the Vilho, Yrjö ja Kalle Väisälän rahasto of the Finnish Academy of Science and Letters, which supported the doctoral studies underlying this work and further acknowledges support for T.K. from the Finnish Cultural Foundation.

The research was conducted using the Estonian Center of Genomics/Roadmap II funded by the Estonian Research Council (project number TT17). This project has received funding from the Estonian Research Council (grants PRG1197 and PRG2625). We acknowledge all the participants of the Estonian Biobank. Data analysis was carried out in part in the High-Performance Computing Center of the University of Tartu.

## References

1. Dowell, D. CDC Clinical Practice Guideline for Prescribing Opioids for Pain — United States, 2022. MMWR Recomm. Rep. 71, (2022).

2. Zanocco, K. et al. Drivers of Variation in Opioid Prescribing after Common Surgical Procedures in a Large Multihospital Healthcare System. J. Am. Coll. Surg. 239, 242–252 (2024).

3. Schieber, L. Z. Variation in Adult Outpatient Opioid Prescription Dispensing by Age and Sex — United States, 2008–2018. MMWR Morb. Mortal. Wkly. Rep. 69, (2020).

4. Koob, G. F. & Volkow, N. D. Neurocircuitry of Addiction. Neuropsychopharmacology 35, 217–238 (2010).

5. Kreek, M. J. et al. Opiate addiction and cocaine addiction: underlying molecular neurobiology and genetics. J. Clin. Invest. 122, 3387–3393 (2012).

6. de Boer, H. D., Detriche, O. & Forget, P. Opioid-related side effects: Postoperative ileus, urinary retention, nausea and vomiting, and shivering. A review of the literature. Best Pract. Res. Clin. Anaesthesiol. 31, 499–504 (2017).

7. Imam, M. Z., Kuo, A., Ghassabian, S. & Smith, M. T. Progress in understanding mechanisms of opioid-induced gastrointestinal adverse effects and respiratory depression. Neuropharmacology 131, 238–255 (2018).

8. Deak, J. D. et al. Genome-wide association study in individuals of European and African ancestry and multi-trait analysis of opioid use disorder identifies 19 independent genome-wide significant risk loci. Mol. Psychiatry 27, 3970–3979 (2022).

9. Mills, S. E. E., Nicolson, K. P. & Smith, B. H. Chronic pain: a review of its epidemiology and associated factors in population-based studies. Br. J. Anaesth. 123, e273–e283 (2019).

10. Vos, T. et al. Global, regional, and national incidence, prevalence, and years lived with disability for 328 diseases and injuries for 195 countries, 1990–2016: a systematic analysis for the Global Burden of Disease Study 2016. The Lancet 390, 1211–1259 (2017).

11. Nicol, A. L., Hurley, R. W. & Benzon, H. T. Alternatives to Opioids in the Pharmacologic Management of Chronic Pain Syndromes: A Narrative Review of Randomized, Controlled, and Blinded Clinical Trials. Anesth. Analg. 125, 1682 (2017).

12. Volkow, N. D. & McLellan, A. T. Opioid Abuse in Chronic Pain — Misconceptions and Mitigation Strategies. N. Engl. J. Med. 374, 1253–1263 (2016).

13. Kolodny, A. et al. The Prescription Opioid and Heroin Crisis: A Public Health Approach to an Epidemic of Addiction. Annu. Rev. Public Health 36, 559–574 (2015).

14. Roberts, E., Copeland, C., Humphreys, K. & Shover, C. L. Drug-related deaths among housed and homeless individuals in the UK and the USA: comparative retrospective cohort study. Br. J. Psychiatry 223, 562–568 (2023).

15. Crews, K. R. et al. Clinical Pharmacogenetics Implementation Consortium Guideline for CYP2D6, OPRM1, and COMT Genotypes and Select Opioid Therapy. Clin. Pharmacol. Ther. 110, 888–896 (2021).

16. Storey, J. D. The positive false discovery rate: a Bayesian interpretation and the q-value. Ann. Stat. 31, 2013–2035 (2003).

17. Toikumo, S. et al. A multi-ancestry genetic study of pain intensity in 598,339 veterans. Nat. Med. 30, 1075–1084 (2024).

18. Walters, R. K. et al. Transancestral GWAS of alcohol dependence reveals common genetic underpinnings with psychiatric disorders. Nat. Neurosci. 21, 1656–1669 (2018).

19. Saunders, G. R. B. et al. Genetic diversity fuels gene discovery for tobacco and alcohol use. Nature 612, 720–724 (2022).

20. Hatoum, A. S. et al. Multivariate genome-wide association meta-analysis of over 1 million subjects identifies loci underlying multiple substance use disorders. Nat. Ment. Health 1, 210–223 (2023).

21. Johnson, E. C. et al. A large-scale genome-wide association study meta-analysis of cannabis use disorder. Lancet Psychiatry 7, 1032–1045 (2020).

22. Toikumo, S. et al. Gene discovery and pleiotropic architecture of Chronic Pain in a Genome-wide Association Study of >1.2 million Individuals. medRxiv 2025.02.28.25323112 (2025) doi:10.1101/2025.02.28.25323112.

23. Kurki, M. I. et al. FinnGen provides genetic insights from a well-phenotyped isolated population. Nature 613, 508–518 (2023).

24. Leitsalu, L., et al. Cohort Profile: Estonian Biobank of the Estonian Genome Center, University of Tartu. Int. J. Epidemiol. 44, 1137–1147 (2015).

25. Bycroft, C. et al. The UK Biobank resource with deep phenotyping and genomic data. Nature 562, 203–209 (2018).

26. Barnett, K. et al. Epidemiology of multimorbidity and implications for health care, research, and medical education: a cross-sectional study. Lancet Lond. Engl. 380, 37–43 (2012).

27. Fayaz, A., Croft, P., Langford, R. M., Donaldson, L. J. & Jones, G. T. Prevalence of chronic pain in the UK: a systematic review and meta-analysis of population studies. BMJ Open 6, e010364 (2016).

28. Okifuji, A. & Hare, B. D. The association between chronic pain and obesity. J. Pain Res. 8, 399–408 (2015).

29. Stone, A. A. & Broderick, J. E. Obesity and pain are associated in the United States. Obes. Silver Spring Md 20, 1491–1495 (2012).

30. Bartley, E. J. & Fillingim, R. B. Sex differences in pain: a brief review of clinical and experimental findings. BJA Br. J. Anaesth. 111, 52–58 (2013).

31. Greenspan, J. D. et al. Studying sex and gender differences in pain and analgesia: A consensus report. Pain 132, S26–S45 (2007).

32. McIntosh, A. M. et al. Genetic and Environmental Risk for Chronic Pain and the Contribution of Risk Variants for Major Depressive Disorder: A Family-Based Mixed-Model Analysis. PLoS Med. 13, e1002090 (2016).

33. Bjornsdottir, G. et al. Rare SLC13A1 variants associate with intervertebral disc disorder highlighting role of sulfate in disc pathology. Nat. Commun. 13, 634 (2022).

34. Liu, S. et al. Spinal apolipoprotein E is involved in inflammatory pain via regulating lipid metabolism and glial activation in the spinal dorsal horn. Biol. Direct 18, 85 (2023).

35. Rivero, O. et al. Cadherin-13, a risk gene for ADHD and comorbid disorders, impacts GABAergic function in hippocampus and cognition. Transl. Psychiatry 5, e655–e655 (2015).

36. Enna, S. J. & McCarson, K. E. The role of GABA in the mediation and perception of pain. Adv. Pharmacol. San Diego Calif 54, 1–27 (2006).

37. Johnston, K. J. A. et al. Genome-wide association study of multisite chronic pain in UK Biobank. PLOS Genet. 15, e1008164 (2019).

38. Mocci, E. et al. Genome wide association joint analysis reveals 99 risk loci for pain susceptibility and pleiotropic relationships with psychiatric, metabolic, and immunological traits. PLOS Genet. 19, e1010977 (2023).

39. Suri, P. et al. Genome-wide meta-analysis of 158,000 individuals of European ancestry identifies three loci associated with chronic back pain. PLoS Genet. 14, e1007601 (2018).

40. Freidin, M. B. et al. Insight into the genetic architecture of back pain and its risk factors from a study of 509,000 individuals. Pain 160, 1361–1373 (2019).

41. Liang, D.-Y. et al. The Netrin-1 receptor DCC is a regulator of maladaptive responses to chronic morphine administration. BMC Genomics 15, 345 (2014).

42. Belonogova, N. M. et al. A multi-trait approach identified 7 novel genes for back pain. Pain Rep. 10, e1218 (2024).

43. Drgon, T. et al. Genome wide association for nicotine dependence and smoking cessation success in NIH research volunteers. Mol. Med. 15, 21–27 (2009).

44. Karlsson Linnér, R., et al. Multivariate analysis of 1.5 million people identifies genetic associations with traits related to self-regulation and addiction. Nat. Neurosci. 24, 1367–1376 (2021).

45. Meng, W. et al. A genome-wide association study finds genetic variants associated with neck or shoulder pain in UK Biobank. Hum. Mol. Genet. 29, 1396–1404 (2020).

46. Liu, L., Liu, M., Song, Z. & Zhang, H. Silencing of FTO inhibits oxidative stress to relieve neuropathic pain by m6A modification of GPR177. Immun. Inflamm. Dis. 12, e1345 (2024).

47. Zhang, F. et al. Inhibition of phosphodiesterase-4 in the spinal dorsal horn ameliorates neuropathic pain via cAMP-cytokine-Cx43 signaling in mice. CNS Neurosci. Ther. 28, 749–760 (2022).

48. Song, W. et al. A genome-wide Association study of the Count of Codeine prescriptions. Sci. Rep. 14, 22780 (2024).

49. Xie, Y., Huo, F. & Tang, J. Cerebral cortex modulation of pain. Acta Pharmacol. Sin. 30, 31–41 (2009).

50. Cao, B. et al. Pathology of pain and its implications for therapeutic interventions. Signal Transduct. Target. Ther. 9, 155 (2024).

51. Kuner, R. & Flor, H. Structural plasticity and reorganisation in chronic pain. Nat. Rev. Neurosci. 18, 20–30 (2017).

52. Bushnell, M. C., Čeko, M. & Low, L. A. Cognitive and emotional control of pain and its disruption in chronic pain. Nat. Rev. Neurosci. 14, 502–511 (2013).

53. Dahl, A. et al. Phenotype integration improves power and preserves specificity in biobank-based genetic studies of major depressive disorder. Nat. Genet. 55, 2082–2093 (2023).

54. An, U. et al. Deep learning-based phenotype imputation on population-scale biobank data increases genetic discoveries. Nat. Genet. 1–8 (2023) doi:10.1038/s41588-023-01558-w.

55. Gu, L.-L. et al. Rapid and accurate multi-phenotype imputation for millions of individuals. Nat. Commun. 16, 387 (2025).

56. Steinfeldt, J. et al. Medical history predicts phenome-wide disease onset and enables the rapid response to emerging health threats. Nat. Commun. 16, 585 (2025).

57. Miao, J. et al. Valid inference for machine learning-assisted genome-wide association studies. Nat. Genet. 56, 2361–2369 (2024).

58. Chen, R. et al. Genetic analyses of eight complex diseases using predicted continuous representations of disease. *Cell Rep*. Methods 5, (2025).

59. Pedersen, E. M. et al. Accounting for age of onset and family history improves power in genome-wide association studies. Am. J. Hum. Genet. 109, 417–432 (2022).

60. Lu, T., Forgetta, V., Richards, J. B. & Greenwood, C. M. T. Capturing additional genetic risk from family history for improved polygenic risk prediction. *Commun*. Biol. 5, 595 (2022).

61. Yang, L., Sadler, M. C. & Altman, R. B. Genetic association studies using disease liabilities from deep neural networks. Am. J. Hum. Genet. 112, 675–692 (2025).

62. Cosentino, J. et al. Inference of chronic obstructive pulmonary disease with deep learning on raw spirograms identifies new genetic loci and improves risk models. Nat. Genet. 55, 787–795 (2023).

63. Forrest, I. S. et al. Machine learning-based marker for coronary artery disease: derivation and validation in two longitudinal cohorts. The Lancet 401, 215–225 (2023).

64. Watanabe, K. et al. Genome-wide meta-analysis of insomnia prioritizes genes associated with metabolic and psychiatric pathways. Nat. Genet. 54, 1125–1132 (2022).

65. Reizine, N. et al. Impact of CYP2D6 Pharmacogenomic Status on Pain Control Among Opioid-Treated Oncology Patients. The Oncologist 26, e2042–e2052 (2021).

66. Dean, L. & Kane, M. Codeine Therapy and CYP2D6 Genotype. in Medical Genetics Summaries (eds Pratt, V. M. et al.) (National Center for Biotechnology Information (US), Bethesda (MD), 2012).

67. Tyndale, R. F., Droll, K. P. & Sellers, E. M. Genetically deficient CYP2D6 metabolism provides protection against oral opiate dependence. Pharmacogenetics 7, 375–379 (1997).

68. Smith, D. M. et al. CYP2D6-guided opioid therapy improves pain control in CYP2D6 intermediate and poor metabolizers: a pragmatic clinical trial. Genet. Med. 21, 1842–1850 (2019).

69. Magarbeh, L., Gorbovskaya, I., Le Foll, B., Jhirad, R. & Müller, D. J. Reviewing pharmacogenetics to advance precision medicine for opioids. Biomed. Pharmacother. 142, 112060 (2021).

70. Machiela, M. J. & Chanock, S. J. LDlink: a web-based application for exploring population-specific haplotype structure and linking correlated alleles of possible functional variants. Bioinformatics 31, 3555–3557 (2015).

71. Zastrozhin, M. et al. Effect of Genetic Polymorphism of the CYP2D6 Gene on the Efficacy and Safety of Fluvoxamine in Major Depressive Disorder. Am. J. Ther. 29, e26 (2022).

72. Kiiskinen, T. et al. Genetic predictors of lifelong medication-use patterns in cardiometabolic diseases. Nat. Med. 29, 209–218 (2023).

73. Front page. Fimea https://www.fimea.fi/documents/542809/838272/13740_Q4.2008_vaihtokelpoiset.txt, https://spc.fimea.fi/indox/nam/html/nam/humspc/0/259390.pdf, https://www.fimea.fi/documents/160140/1187305/GS-ladattava.txt, https://www.fimea.fi/documents/160140/744738/29472_Varastointivelvoitteen_piriin_kuuluvat_laakevalmisteet_2015-05-19.txt, https://www.fimea.fi/documents/160140/744738/27145_Varastointivelvoitteen_piriin_kuuluvat_laakevalmisteet_2014-08-25.txt.

74. Forside - www.medicinpriser.dk. medicinepricer.dk https://www.medicinpriser.dk/Default.aspx?id=15&vnr=013628.

75. Icelandic Medicine Price Catalogue. Icelandic Medicines Agency https://www.ima.is/home/pricing-and-reimbursement/english-description-of-the-icelandic-medicine-price-catalogue/.

76. admin. Human medicines - Czech Republic/EU. SÚKL http://www.sukl.cz/file/88816_1_1/download/ (2008).

77. Nielsen, S., Degenhardt, L., Hoban, B. & Gisev, N. Comparing opioids: A guide to estimating oral morphine equivalents (OME) in research.

78. Opioid Equivalence Chart. Gloucestershire Hospitals NHS Foundation Trust https://www.gloshospitals.nhs.uk/healthcare-professionals/treatment-guidelines/opioid-equivalence-chart/.

79. Mbatchou, J. et al. Computationally efficient whole-genome regression for quantitative and binary traits. Nat. Genet. 53, 1097–1103 (2021).

80. Li, X., Wong, W., Lamoureux, E. L. & Wong, T. Y. Are Linear Regression Techniques Appropriate for Analysis When the Dependent (Outcome) Variable Is Not Normally Distributed? Invest. Ophthalmol. Vis. Sci. 53, 3082–3083 (2012).

81. Lumley, T., Diehr, P., Emerson, S. & Chen, L. The Importance of the Normality Assumption in Large Public Health Data Sets. Annu. Rev. Public Health 23, 151–169 (2002).

82. Willer, C. J., Li, Y. & Abecasis, G. R. METAL: fast and efficient meta-analysis of genomewide association scans. Bioinformatics 26, 2190–2191 (2010).

83. Hinrichs, A. S. et al. The UCSC Genome Browser Database: update 2006. Nucleic Acids Res. 34, D590–D598 (2006).

84. Wang, G., Sarkar, A., Carbonetto, P. & Stephens, M. A Simple New Approach to Variable Selection in Regression, with Application to Genetic Fine Mapping. J. R. Stat. Soc. Ser. B Stat. Methodol. 82, 1273–1300 (2020).

85. Benner, C. et al. Prospects of Fine-Mapping Trait-Associated Genomic Regions by Using Summary Statistics from Genome-wide Association Studies. Am. J. Hum. Genet. 101, 539–551 (2017).

86. Bulik-Sullivan, B. K. et al. LD Score regression distinguishes confounding from polygenicity in genome-wide association studies. Nat. Genet. 47, 291–295 (2015).

87. Auton, A. et al. A global reference for human genetic variation. Nature 526, 68–74 (2015).

88. Altshuler, D. M. et al. Integrating common and rare genetic variation in diverse human populations. Nature 467, 52–58 (2010).

89. Finucane, H. K. et al. Partitioning heritability by functional annotation using genome-wide association summary statistics. Nat. Genet. 47, 1228–1235 (2015).

90. Finucane, H. K. et al. Heritability enrichment of specifically expressed genes identifies disease-relevant tissues and cell types. Nat. Genet. 50, 621–629 (2018).

91. GTEx Consortium. The GTEx Consortium atlas of genetic regulatory effects across human tissues. Science 369, 1318–1330 (2020).

92. Open Targets Platform: facilitating therapeutic hypotheses building in drug discovery | Nucleic Acids Research | Oxford Academic. https://academic.oup.com/nar/article/53/D1/D1467/7917960?login=false.

93. Ghoussaini, M. et al. Open Targets Genetics: systematic identification of trait-associated genes using large-scale genetics and functional genomics. Nucleic Acids Res. 49, D1311–D1320 (2021).

94. Chen, T. & Guestrin, C. XGBoost: A Scalable Tree Boosting System. in *Proceedings of the 22nd ACM SIGKDD International Conference on Knowledge Discovery and Data Mining* 785–794 (Association for Computing Machinery, New York, NY, USA, 2016). doi:10.1145/2939672.2939785.

