## Supplement-Material for "Machine learning augmented genome-wide meta-analysis of prescription opioid use in 860,000 individuals"

### FinnGen Study Cohort and Genetic Data

#### Cohort Overview

FinnGen is a nationwide research project that integrates genotype data from over 520,000 Finnish biobank participants with longitudinal national health register data. The cohort includes individuals recruited through hospital biobanks, disease-specific studies, and blood donor registries, with intentional enrichment for disease cases to enhance statistical power for genetic discovery. FinnGen participants had a median age of 53 years at sample collection, with 57% female and 43% male.

#### Genotyping and Quality Control

Participants were genotyped using custom ThermoFisher Axiom arrays: 121,000 samples on version 1 (657,675 markers) and ~328,000 on version 2 (723,376 probe sets for 664,510 markers). Arrays include GWAS backbones and variants relevant to coding regions, pharmacogenomics, immune traits, and Finnish-enriched alleles.

Sample-level QC excluded individuals based on:

- Sex discordance between genetically inferred and registry-reported sex (F-statistic $< 0.4$ for females, $> 0.7$ for males),
- Genotype missingness $> 2\%$,
- Excess heterozygosity in common variants (allele frequency $> 0.05$, $> 3$ SD from batch mean),
- Excess relatedness ($\hat{\pi} > 0.1$), removed in two rounds: first for samples related to $> 500$ others, then for those related to $> 50$ others.

Variant-level QC removed markers that:

- Had non-ATCG alleles,
- Were absent from the SISu v4.2 imputation panel or had panel allele frequency $< 0.001$ (imputation only),
- Showed significant allele frequency deviation from the panel ($p < 5\times10^{-8}$, adjusted for 10 PCs),
- Had Hardy-Weinberg disequilibrium p-value $< 1\times10^{-10}$ across all batches (with exceptions for rare variants with homozygote deficiency),
- Were missing in $> 15\%$ of batches or non-PASS in $> 30\%$ of batches.

#### Imputation and Ancestry Assignment

Genotyped samples were pre-phased with Eagle 2.3.5 using 20,000 conditioning haplotypes and imputed using the Finnish-specific SISu v4.2 reference panel. The panel was built from high-coverage (25x) whole-genome sequencing of 8,554 Finnish individuals and mapped to GRCh38.

Ancestry was implicitly accounted for by using a Finnish-specific reference panel, eliminating the need for global ancestry prediction. Population outliers and samples with excessive relatedness were removed as part of quality control to reduce cryptic structure.

#### Phenotype Definition from Health Registers

FinnGen uses harmonized clinical phenotype definitions based on structured data from a wide range of nationwide health and social care registers. The following data sources were used:

- Statistics Finland
- Finnish Cancer Registry and Mass Screening Registry
- Register of Primary Health Care Visits (Avohilmo)
- Care Register for Health Care
- The Social Insurance Institution of Finland (Kela)
- Digital and Population Data Services Agency
- Finnish Register of Visual Impairment
- Care Register for Social Welfare
- Finnish Registry for Kidney Diseases
- Finnish National Infectious Diseases Register
- Medical Birth Register
- Finnish National Vaccination Register
- Register of Congenital Malformations

These registers provide detailed information on diagnoses, procedures, prescriptions, hospitalizations, and mortality. Phenotypes were defined using harmonized algorithms developed by expert working groups and are publicly available. This study used definitions from FinnGen Data Freeze 12 (DF12).

#### UK Biobank Study Cohort and Genetic Data

**Cohort Overview**
UK Biobank is a population-based prospective cohort of approximately 500,000 individuals aged 40–69 at recruitment between 2006 and 2010. Participants provided biological samples and consent for long-term follow-up via linkage to national health records.

**Genotyping and Quality Control**
Participants were genotyped using two closely related arrays: 49,950 individuals with the UK BiLEVE Axiom array and ~438,000 with the UK Biobank Axiom array. Both arrays share >95% marker content and include ~800,000 variants selected for genome-wide imputation coverage and trait relevance. Genotyping was performed at the Affymetrix Research Services Laboratory in batches of ~4,800 samples.

Variant-level QC was applied batch-wise using Affymetrix protocols and custom filters. Variants were excluded if they exhibited:

- Hardy-Weinberg disequilibrium ($p < 1\times10^{-12}$),
- Plate or batch effects ($p < 1\times10^{-12}$ via Fisher's exact test),
- 2% missingness across samples,
- Poor clustering or discordance in replicate control samples.

Samples were excluded for:

- Sex mismatches (self-reported vs. genetic),
- Excess heterozygosity or missingness (adjusted for population structure),
- Sex chromosome aneuploidy,
- Relatedness or duplicates, if unresolved.

**Imputation and Ancestry Assignment**
Imputation was performed for 487,442 individuals using IMPUTE4 with a combined reference panel from the Haplotype Reference Consortium (HRC) and UK10K + 1000 Genomes. Variants were retained only if present in both panels, resulting in ~97 million imputed SNPs.

Ancestry inference used PCA on a curated set of ~467,000 autosomal SNPs shared with HapMap3 and present in the imputed data. Individuals were assigned to five super-populations (AFR, AMR, EAS, EUR, SAS) using GenoPred's multinomial elastic-net classifier trained on 1000 Genomes data. Outliers were excluded based on Mahalanobis distance from ancestry-specific centroids.

**Phenotype Definition from Health Registers**
UK Biobank provides linked health data including hospital inpatient diagnoses (ICD-9/10, OPCS-3/4), death records (ICD-10), primary care data (READ2, CTV-3), and cancer registry entries. Phenotypes were derived using first occurrence mappings and algorithmically-defined outcomes curated from structured health records. Censoring dates for hospital, cancer, and death data were set at 31 January 2021.

#### Estonian Biobank Study Cohort and Genetic Data

**Cohort Overview**
The Estonian Biobank (EstBB)[1] is a population-based biobank managed by the Institute of Genomics at the University of Tartu. The cohort includes over 200,000 participants, representing approximately 20% of Estonia's adult population. Participants have consented to long-term follow-up through linkage to national electronic health records, enabling detailed longitudinal phenotyping.

**Genotyping and Quality Control**
Genotyping was performed at the University of Tartu's Core Genotyping Lab using four sub-versions of the Illumina Global Screening Array (GSAv1.0, GSAv2.0, GSAv2.0_EST, and GSAv3.0). In total, 212,955 DNA samples were genotyped.

Samples were excluded for:

- Call rate < 95%,
- Sex mismatch between registry-reported and genetically inferred sex, after manual review for sex chromosome abnormalities.

Following sample QC, 211,259 individuals remained for analysis.

Variants were excluded if they:

- Had call rate < 95%,
- Had HWE p-value < $1\times10^{-4}$ (autosomal SNPs),
- Had Illumina GenTrain score < 0.6 or cluster separation score < 0.4 in any batch,
- Had inconsistent allele frequency among genotyping experiments

All variants were mapped to GRCh38,  Per-variant QC excluded AT/GC SNPs (strand-ambiguous), indels, CNVs, and invariant sites (MAC=0). After QC, 295,935 autosomal and X-chromosome variants were retained.

**Imputation and Ancestry Assignment**
Genotype data were pre-phased with Eagle v2.4.1 (using --Kpbwt=20000) and imputed using Beagle 5.4 (beagle.22Jul22.46e.jar)  with an effective population size of 20,000. Imputation used a population-specific reference panel of 2,695 Estonian whole-genome sequenced individuals.

Ancestry proportions and group assignments were estimated with bigsnpr. For inference, genotypes were imputed using reference haplotypes from the 1000 Genomes Project (N = 2,495) obtained from the HRC panel. In total, ~4.54 million SNPs were used for ancestry estimation.

**Phenotype Definition from Health Registers**
Phenotypic information was derived by linking biobank participants to Estonia's national health insurance fund drug prescription and purchase records from 2004-2023.

**GWAS**
Participants without genotype data or with non-european ancestry, heterozygosity outliers for common SNPs (±3SD) and one random individual per monozygous twins were excluded. Analyses were adjusted for sex, age at death or end of follow-up, age$^2$ and the first ten genetic principal components.

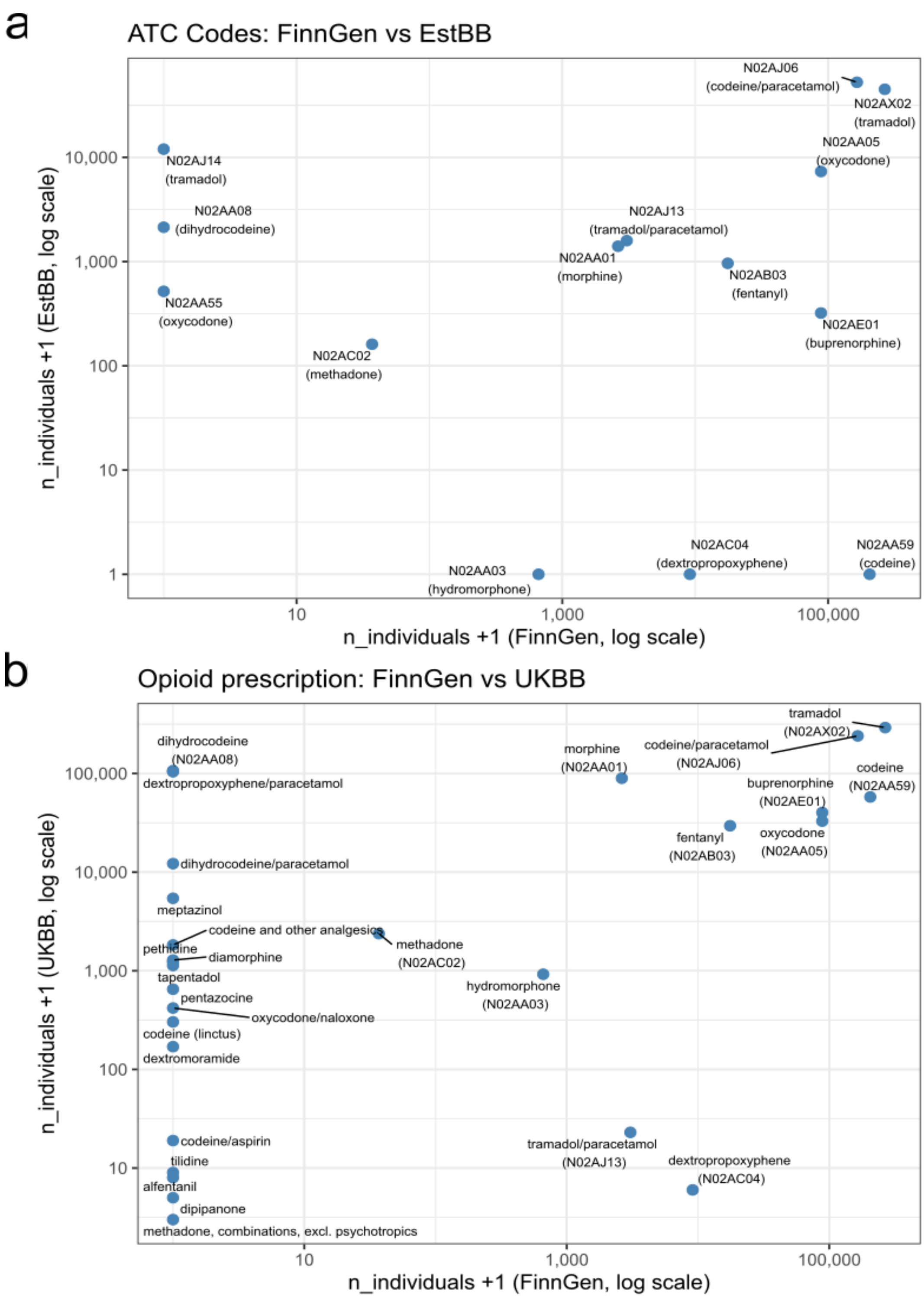


**Supplementary Figure 1 | Prescription counts across biobanks.**

(a) Scatterplot of the number of individuals prescribed each medication in FinnGen (x-axis) and EstBB (y-axis), displayed on a log scale. Each point represents one medication, illustrating strong cross-biobank consistency in prescription frequencies. (b) Scatterplot of prescription counts for the same medications in FinnGen (x-axis) and UKBB (y-axis), also shown on a log scale.

Points with a value of 1 on either axis indicate medications with no recorded prescriptions or medications excluded in that biobank due to low counts required for participant anonymisation.

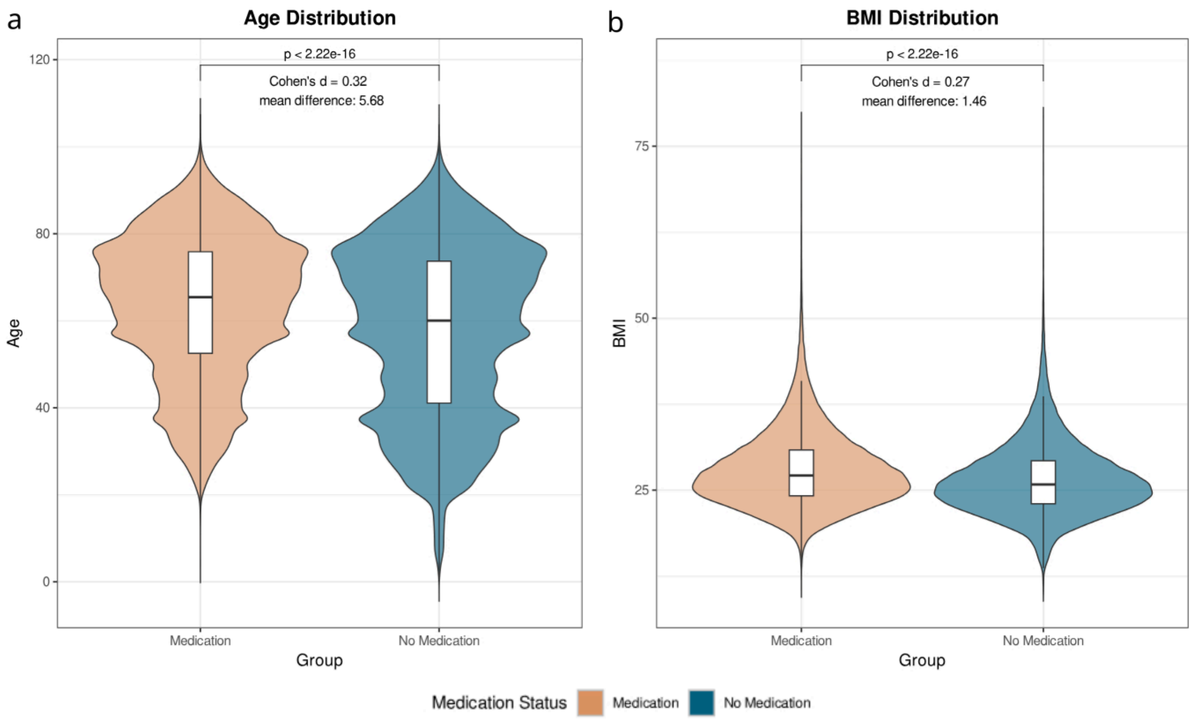


**Supplementary Figure 2 | Population characteristics of individuals with and without opioid prescriptions in FinnGen**

(a) Age distribution, showing older age among individuals with opioid prescriptions (Cohen's *d* = 0.32, mean difference = 5.58 years).

(b) BMI distribution, showing slightly higher BMI among opioid users (Cohen's *d* = 0.27, mean difference = 1.46).

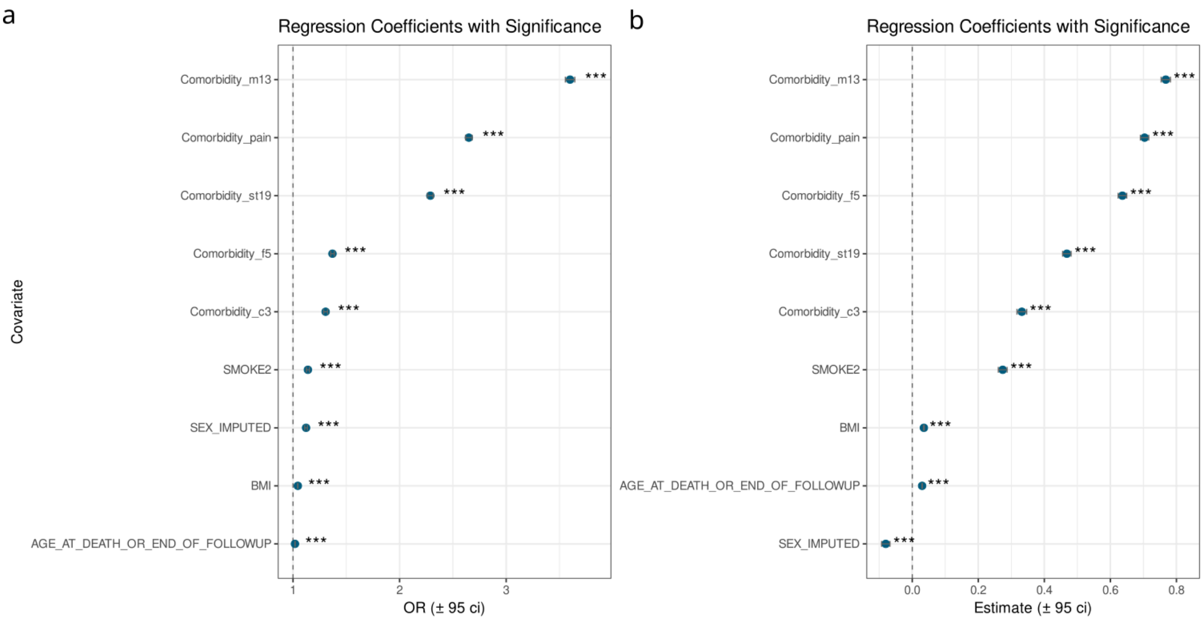


**Supplementary Figure 3 | Clinical and demographic correlates of opioid prescriptions in FinnGen.**

(a) Logistic regression for opioid use (ever vs. never). Opioid prescriptions were most strongly associated with musculoskeletal disorders (M13; OR = 3.60), pain diagnoses (pain; OR = 2.65), and soft-tissue conditions (ST19; OR = 2.29). Psychiatric diagnoses (F5; OR = 1.37), cancer (C3; OR = 1.31), smoking, higher BMI, and older age were also positively associated.

(b) Linear regression among opioid users. Higher prescription counts were similarly driven by M13 (β = 0.77), pain (β = 0.70), F5 (β = 0.64), and ST19 (β = 0.47), with smaller positive effects for C3, smoking, and BMI, and a slight negative association for sex.

FinnGen endpoint abbreviations: F5 = mental and behavioural disorders; M13 = musculoskeletal disorders; C3 = neoplasms; ST19 = soft-tissue/injury-related disorders.

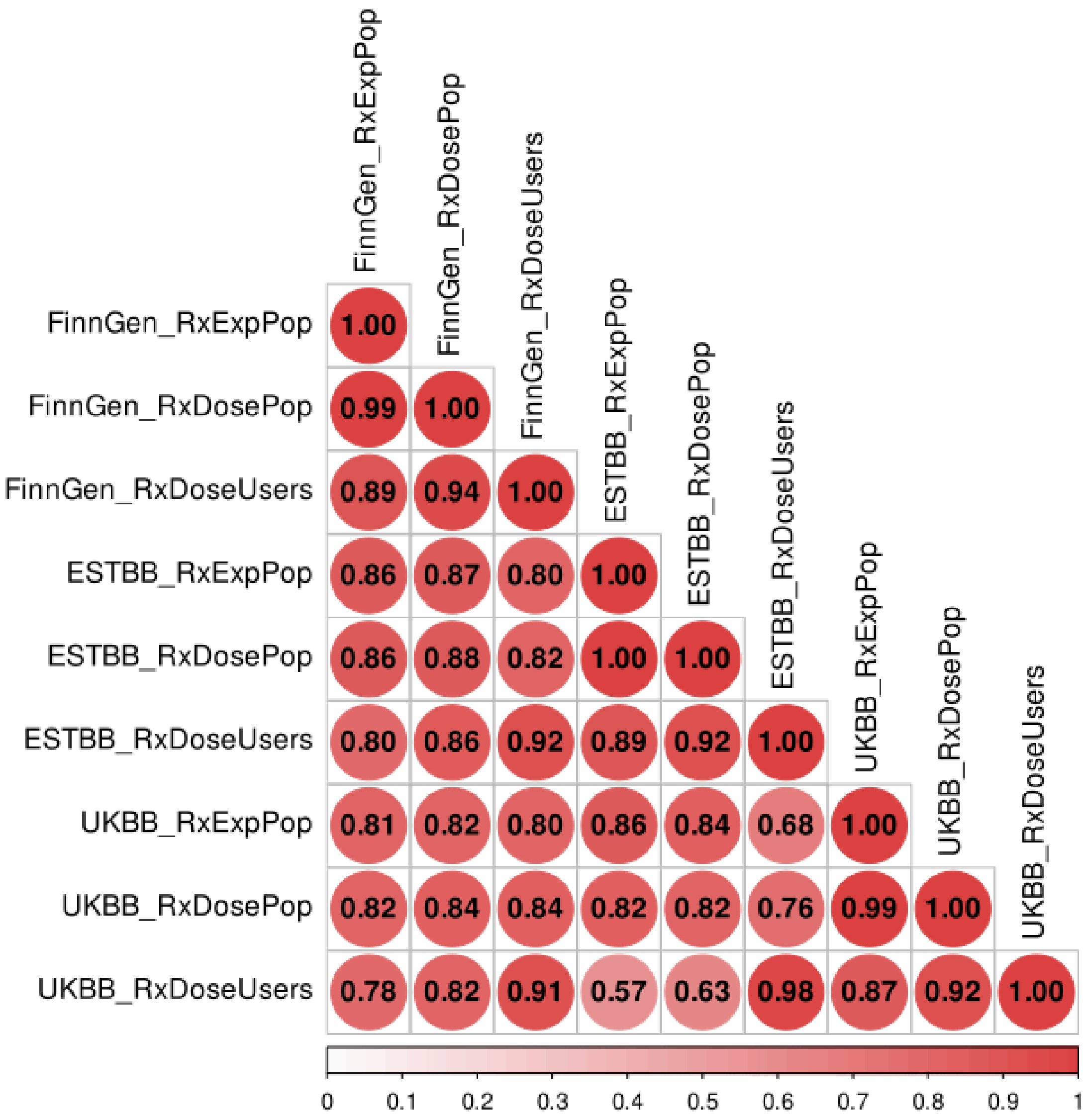

**Supplementary Figure 4 | Genetic correlations of opioid prescription phenotypes across biobanks.**

Genetic correlations (*rg*) were estimated across FinnGen, EstBB, and UK Biobank for the three opioid prescription phenotypes: RxExpPop (binary prescription status), RxDoseUsers (continuous dosage among users), and RxDosePop (combined dosage plus binary status).

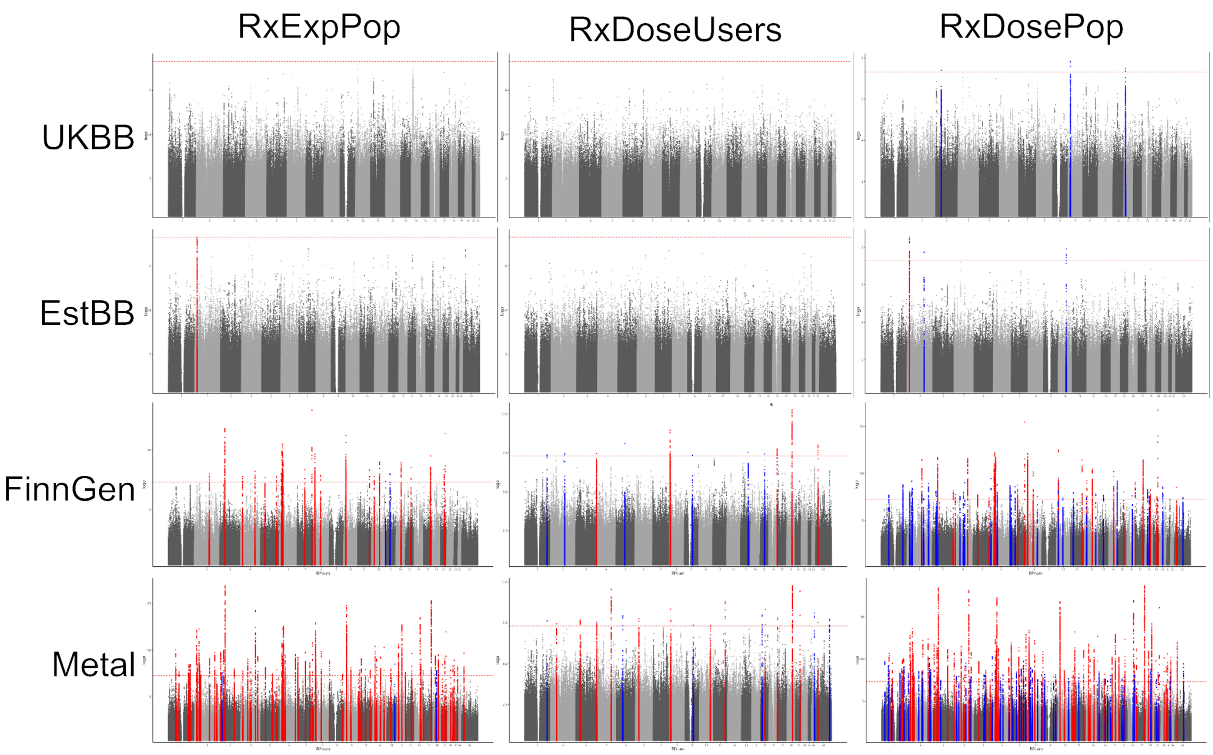


**Supplementary Figure 5 | Genome-wide association results for opioid prescription phenotypes across biobanks.**

Shown are Manhattan plots for the three opioid prescription phenotypes: (a) RxExpPop (binary indicator of ever having an opioid prescription), (b) RxDoseUsers (continuous dosage phenotype among individuals with at least one opioid prescription), and (c) RxDosePop (combined dosage plus binary opioid prescription). Results are displayed for individual biobanks (FinnGen, EstBB, UK Biobank), for within-biobank analyses, and for meta-analyses across cohorts. The red line denotes the genome-wide significance threshold ($P < 5 \times 10^{-8}$).Loci in blue are unique to the trait and loci marked in red are shared with at least one other trait.

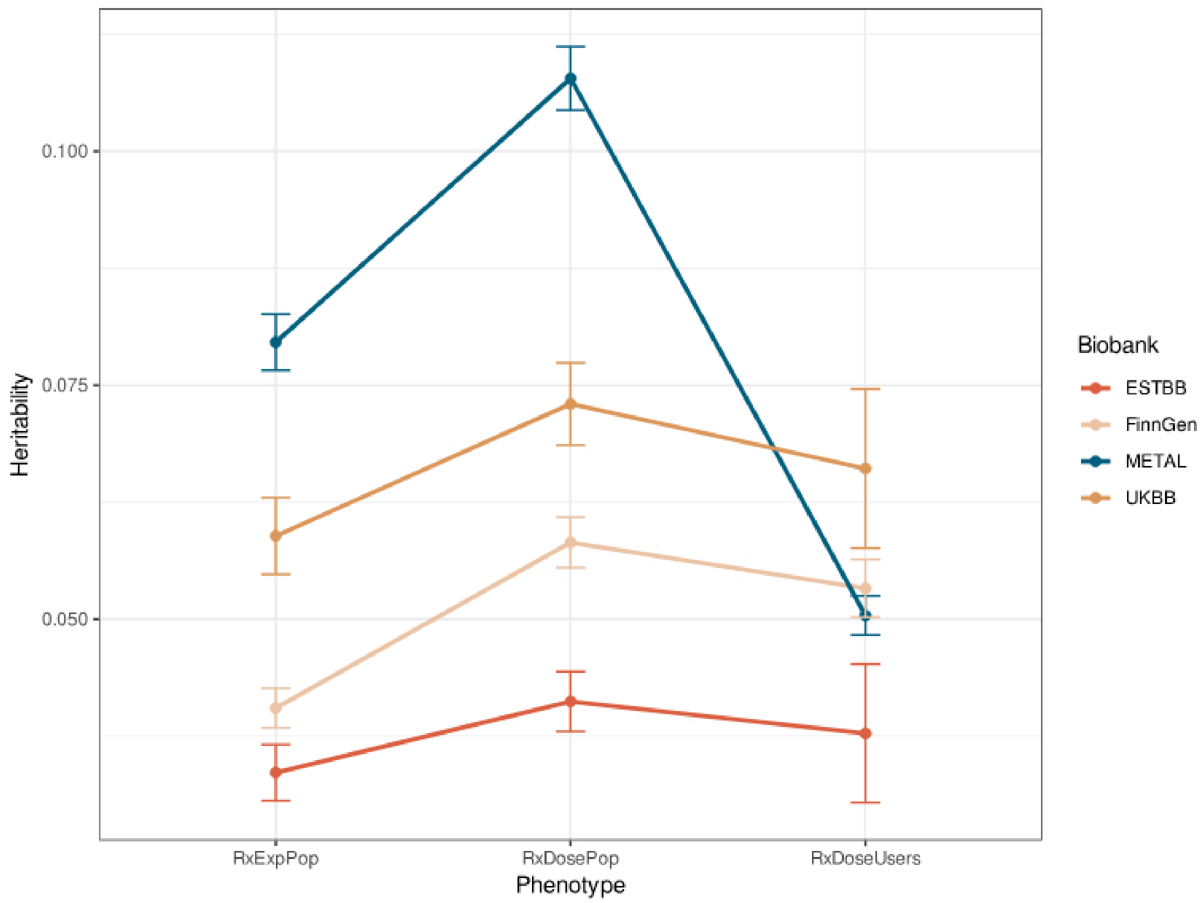


**Supplementary Figure 6 | SNP-based heritability estimates of opioid prescription phenotypes.**

SNP-based heritability ($h^2$_SNP) was estimated for the three opioid prescription phenotypes: RxExpPop (binary ever-prescribed), RxDoseUsers (continuous dosage among users), and RxDosePop (combined dosage plus binary status). Estimates were derived using LD Score Regression. Due to differences in phenotype scale (two continuous traits and one binary trait), direct comparison of heritability values should be interpreted with caution.

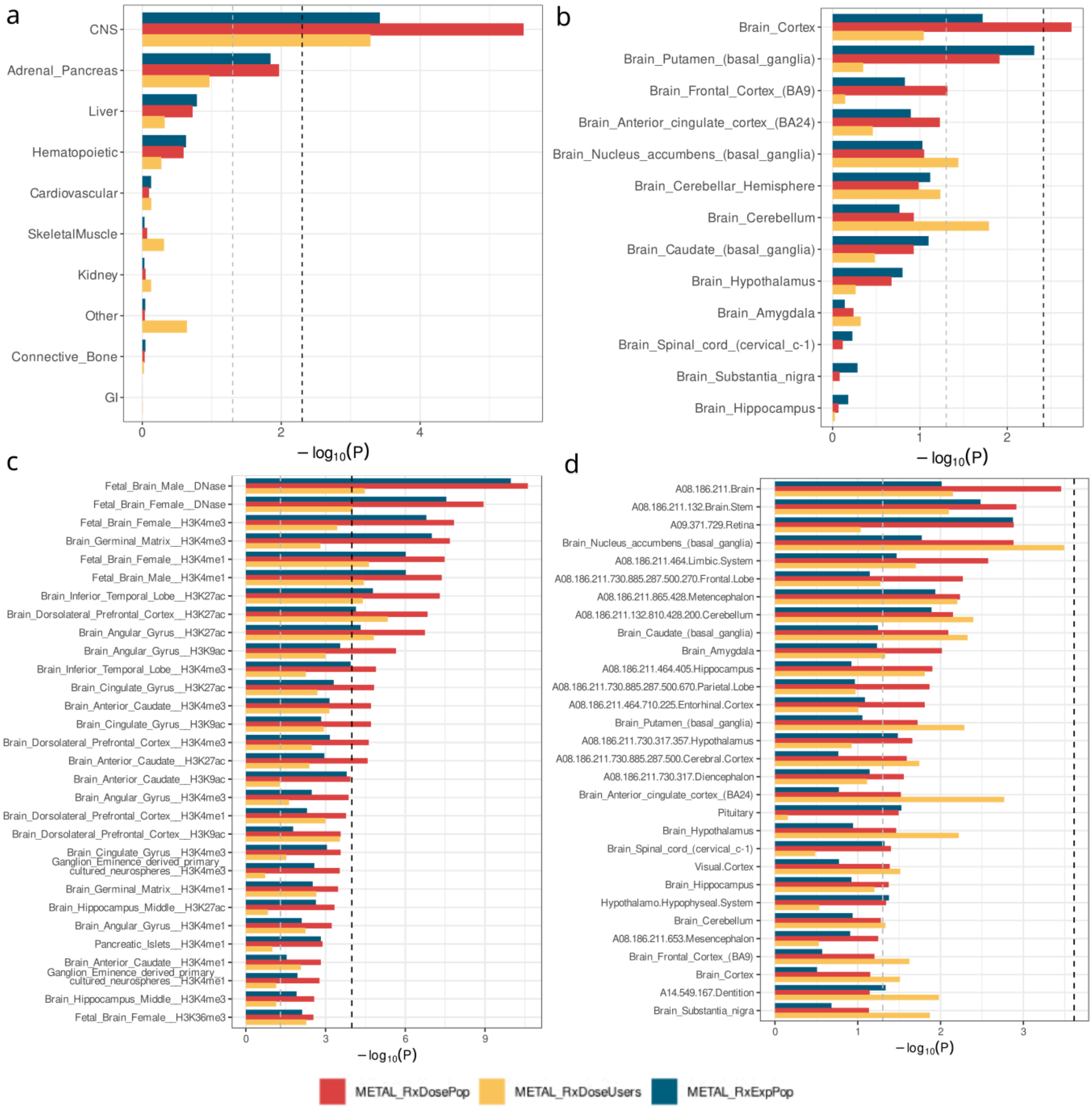


**Supplementary Figure 7 | Partitioned SNP-based heritability of opioid prescription phenotypes.** Partitioned heritability analyses were performed using LD Score Regression to test enrichment of SNP-based heritability ($h^2$_SNP) across tissues and functional annotations. Results are shown for (a) baseline cell type groups, (b) baseline brain regions, (c) tissue-specific chromatin annotations, and (d) additional brain- and tissue-specific annotations. Analyses were conducted separately for the three opioid phenotypes: RxExpPop (binary ever-prescribed), RxDoseUsers (continuous dosage among users), and RxDosePop (combined dosage plus binary status).

Baseline annotation models were derived from the LD Score Regression framework (Finucane et al. 2015; Gazal et al. 2017) and extended with tissue- and chromatin-specific annotations introduced in Finucane et al. 2018 and subsequent updates.

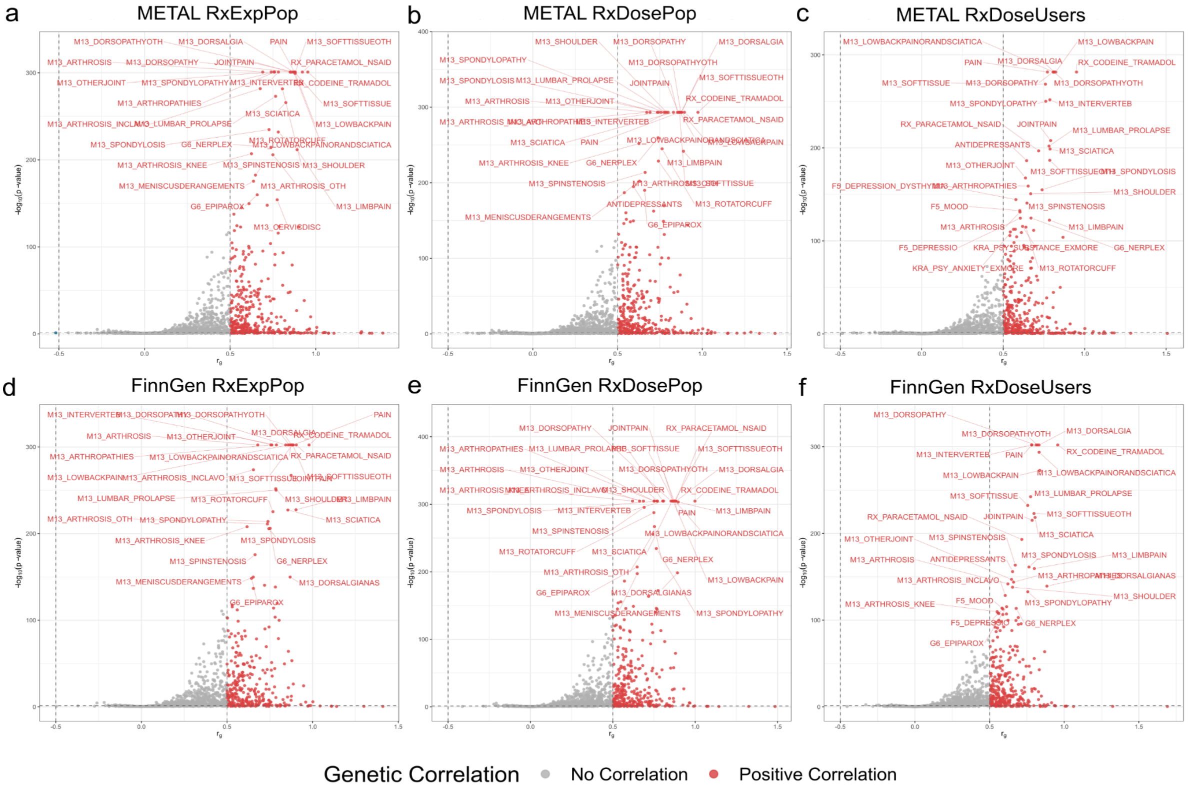


**Supplementary Figure 8 | Genetic correlations of opioid prescription phenotypes with 2,481 FinnGen traits.**

Genetic correlations (*rg*) were estimated using LD Score Regression for (a, d) RxExpPop, (b, e) RxDosePop, and (c, f) RxDoseUsers. Panels (a–c) show results from the METAL meta-analysis, and panels (d–f) from FinnGen-only analyses. Manhattan-style volcano plots display the top 30 significant correlations, primarily with musculoskeletal, pain, and medication-related traits. Multiple testing was controlled using FDR-adjusted *q*-values (Storey & Tibshirani, 2003).

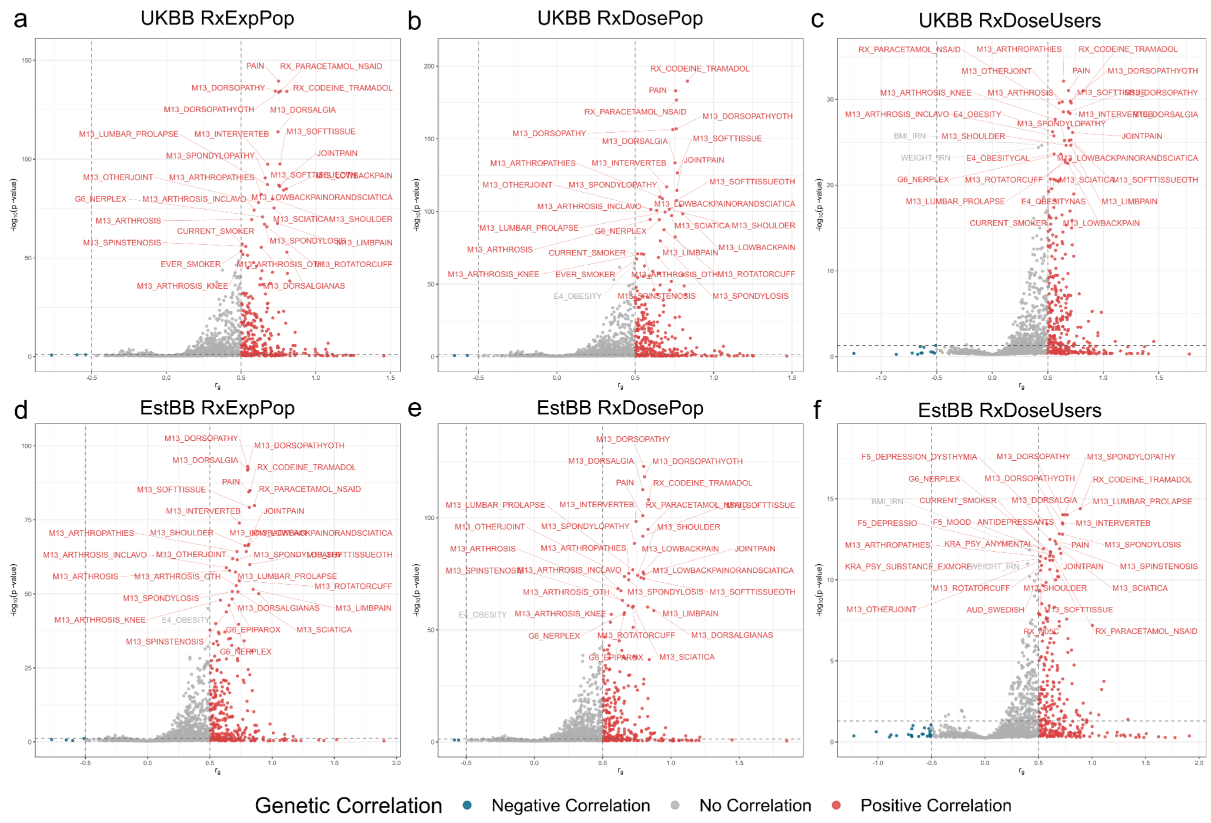


**Supplementary Figure 9 | Genetic correlations of opioid prescription phenotypes with 2,481 FinnGen traits.**

Genetic correlations (*rg*) were estimated using LD Score Regression for (a, d) RxExpPop, (b, e) RxDosePop, and (c, f) RxDoseUsers. Panels (a–c) show results from the UKBB meta-analysis, and panels (d–f) from EstBB analyses. Manhattan-style volcano plots display the top 30 significant correlations, primarily with musculoskeletal, pain, and medication-related traits. Multiple testing was controlled using FDR-adjusted *q*-values (Storey & Tibshirani, 2003).

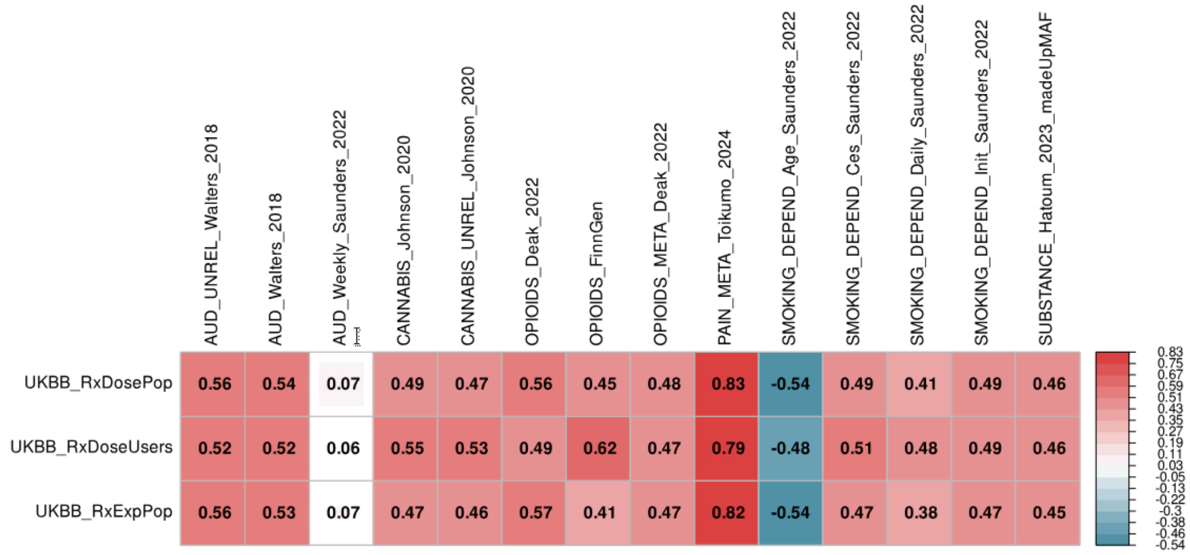

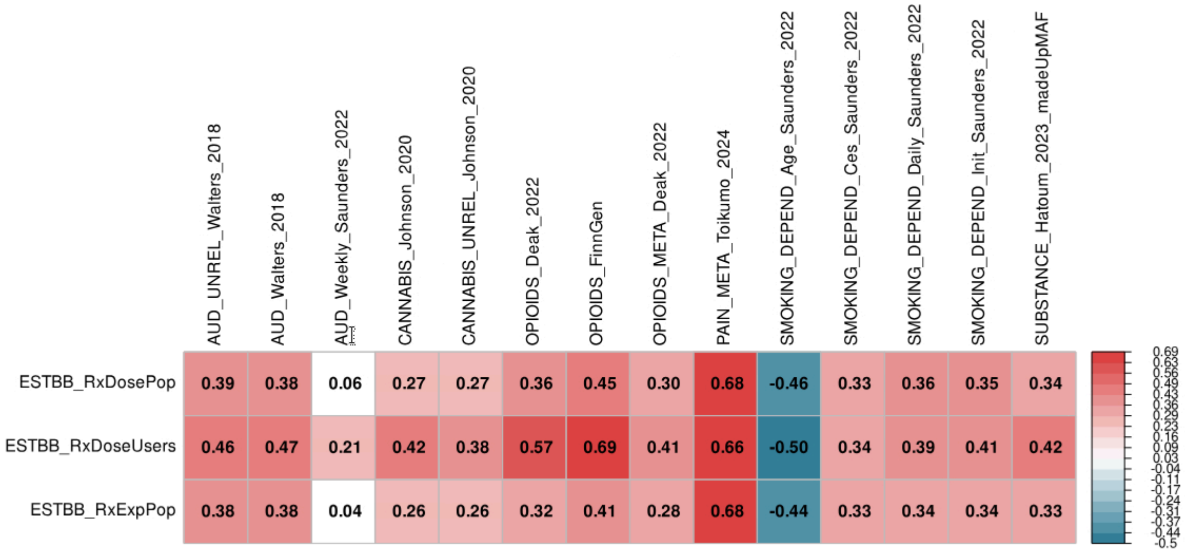


**Supplementary Figure 10 | Genetic correlations of opioid prescription phenotypes in EstBB and UK Biobank with external studies.**

Genetic correlations (*rg*) were estimated using LD Score Regression between opioid prescription phenotypes (RxExpPop, RxDosePop, RxDoseUsers) and published studies of alcohol use disorder[2,3], cannabis use[4], opioid dependence[5], pain[6], smoking behaviors[3], and substance use[7]. (a) EstBB results. (b) UK Biobank results.

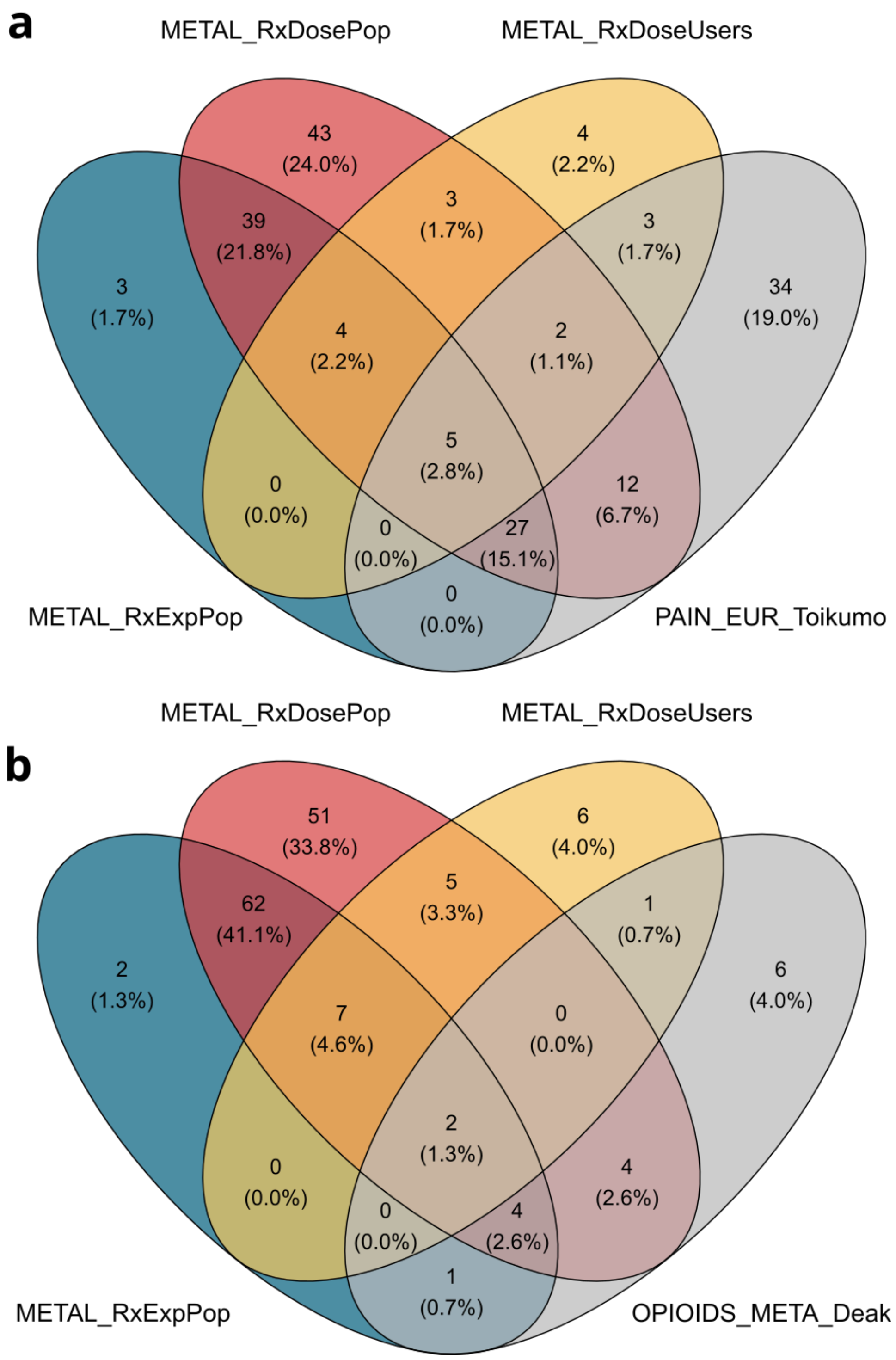


**Supplementary Figure 11 | Lead SNP overlap of opioid prescription phenotypes with external GWAS.**

(a) Overlap of independent lead SNPs from opioid prescription phenotypes (RxExpPop, RxDosePop, RxDoseUsers) with the European GWAS of pain (Tokumo et al. 2024).

(b) Overlap of independent lead SNPs from the same phenotypes with a GWAS of opioid addiction (Deak et al. 2022). Percentages indicate the proportion of overlapping SNPs relative to the total in each study.

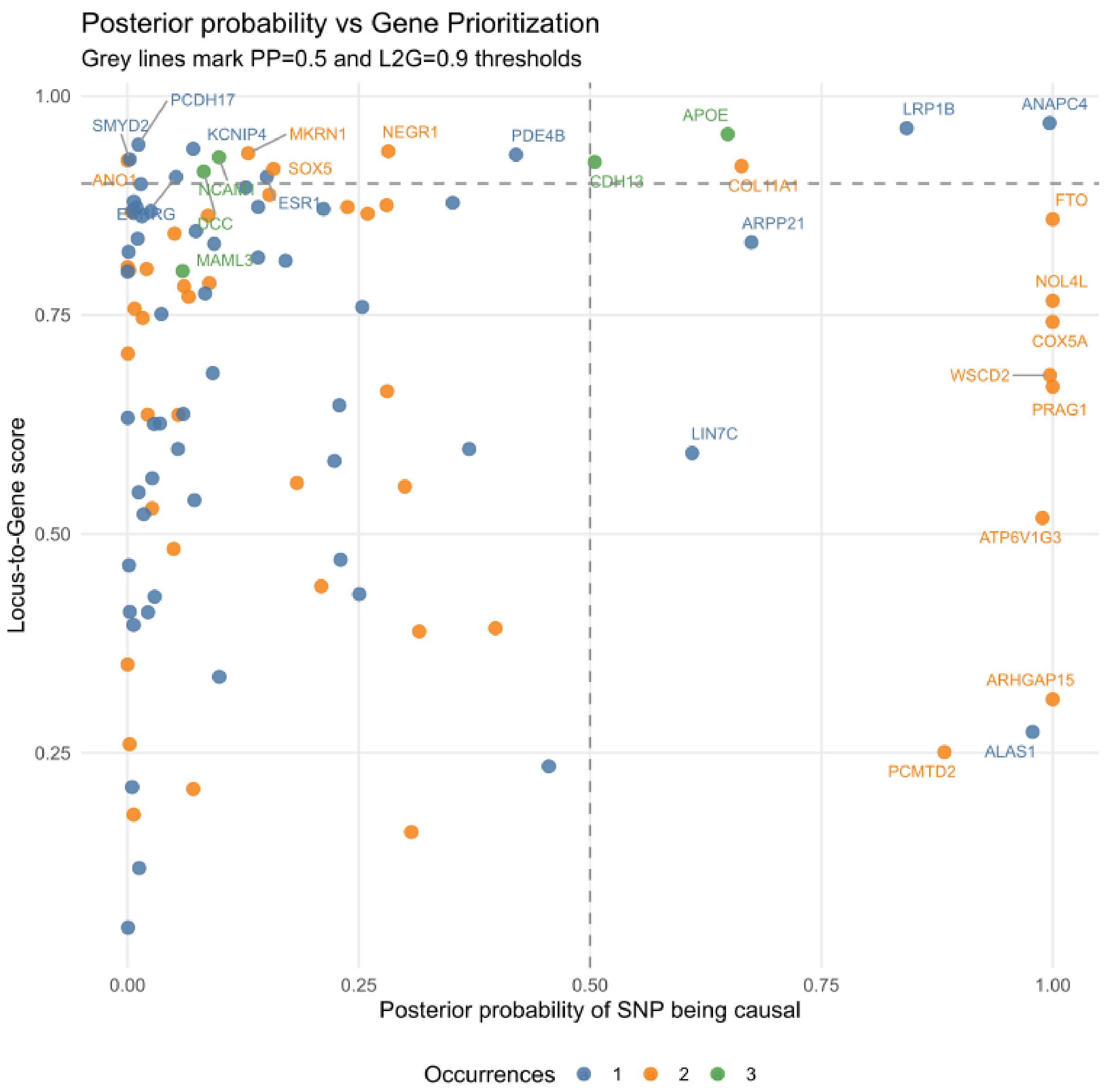


**Supplementary Figure 12 | Gene prioritization using Open Targets Genetics.**

Lead SNPs from opioid prescription GWAS were mapped to candidate genes using posterior probabilities from fine-mapped credible sets and the Open Targets Locus-to-Gene (L2G) score. The plot compares posterior probabilities of SNPs within credible sets against L2G gene-level scores, integrating functional and statistical evidence to highlight likely causal genes.

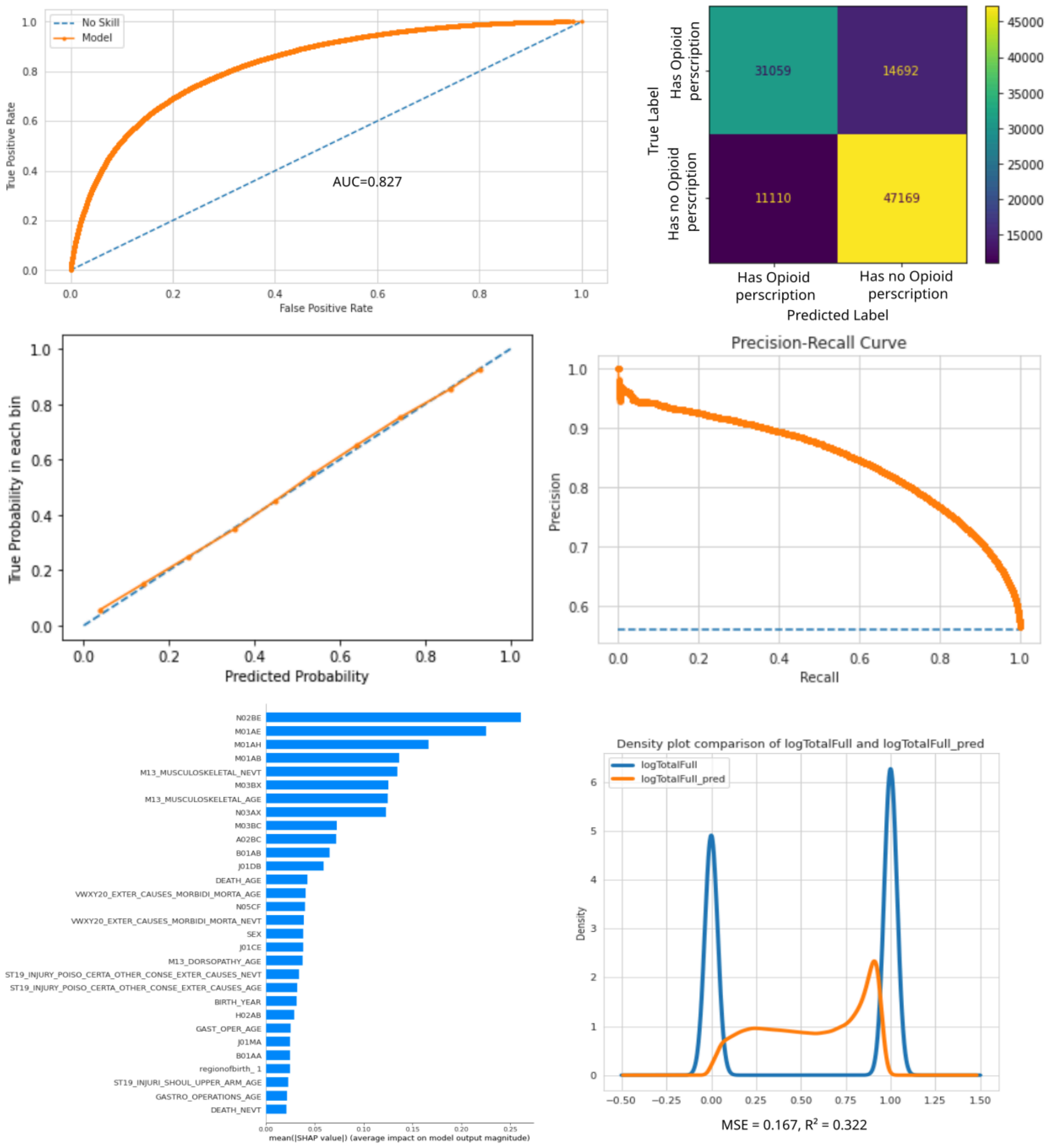


**Supplementary Figure 13 | Performance of the XGBoost model predicting opioid prescription status Model I.**

(a) Receiver operating characteristic (ROC) curve with area under the curve (AUC = 0.827).
(b) Confusion matrix showing 31,059 true positives, 47,169 true negatives, 14,692 false positives, and 11,110 false negatives.
(c) Calibration plot, demonstrating close alignment between predicted and observed probabilities.
(d) Precision-recall curve, with precision >0.6 across most recall values.
(e) Top predictors identified using SHAP values, including musculoskeletal (M13), dorsopathy (M13), and injury-related codes.
(f) Density comparison of observed (blue) and predicted (orange) distributions of binary RxExpPop, with mean squared error (MSE = 0.167) and $R^2$ = 0.322.

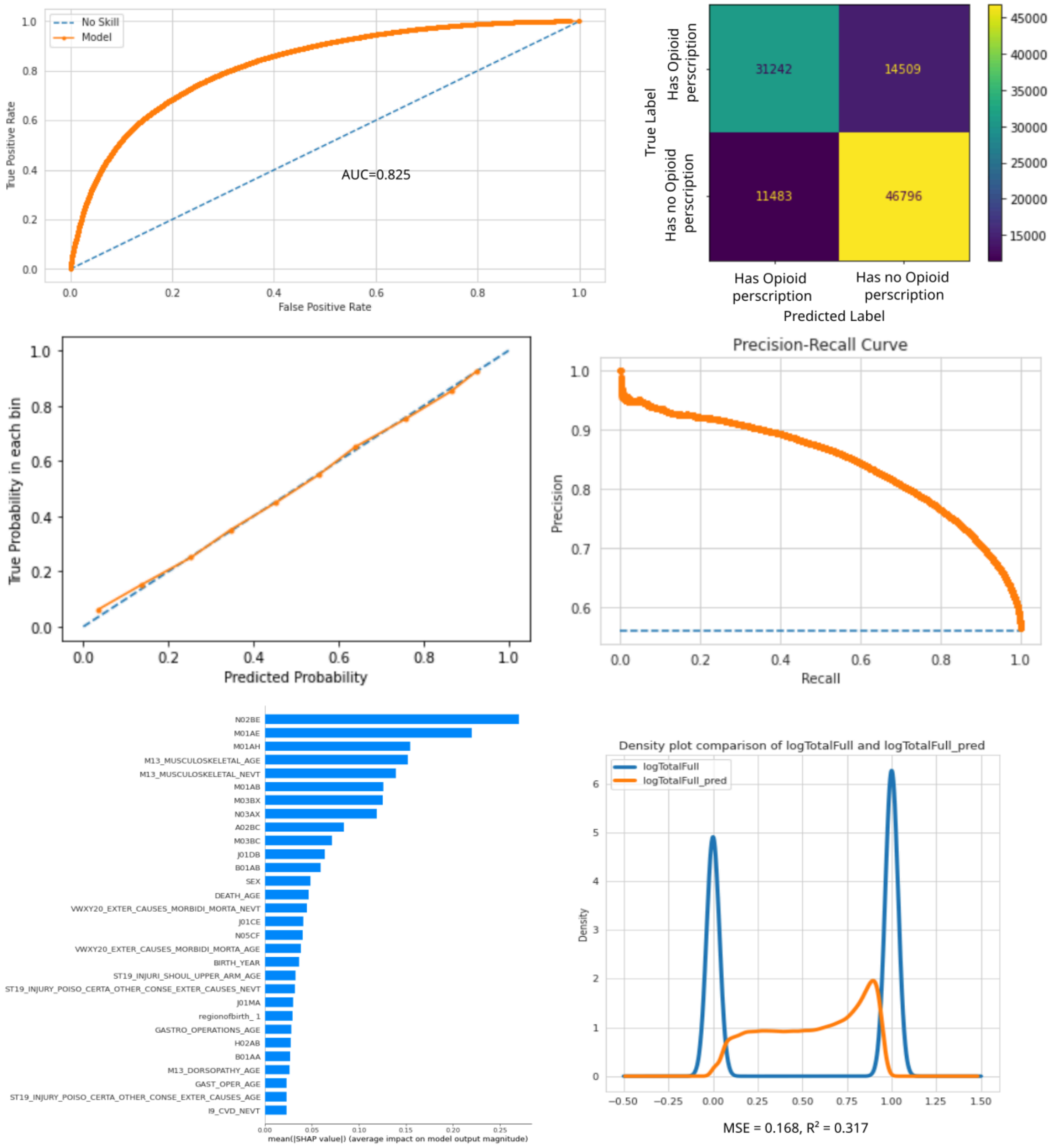


**Supplementary Figure 14 | Performance of the XGBoost model predicting opioid prescription status Model II.**

(a) Receiver operating characteristic (ROC) curve with area under the curve (AUC = 0.825).
(b) Confusion matrix showing 31,242 true positives, 46,796 true negatives, 14,509 false positives, and 11,483 false negatives.
(c) Calibration plot, demonstrating close alignment between predicted and observed probabilities.
(d) Precision-recall curve, with precision >0.6 across most recall values.
(e) Top predictors identified using SHAP values, including musculoskeletal (M13), dorsopathy (M13), and injury-related codes.
(f) Density comparison of observed (blue) and predicted (orange) distributions of binary RxExpPop, with mean squared error (MSE = 0.168) and $R^2$ = 0.317.

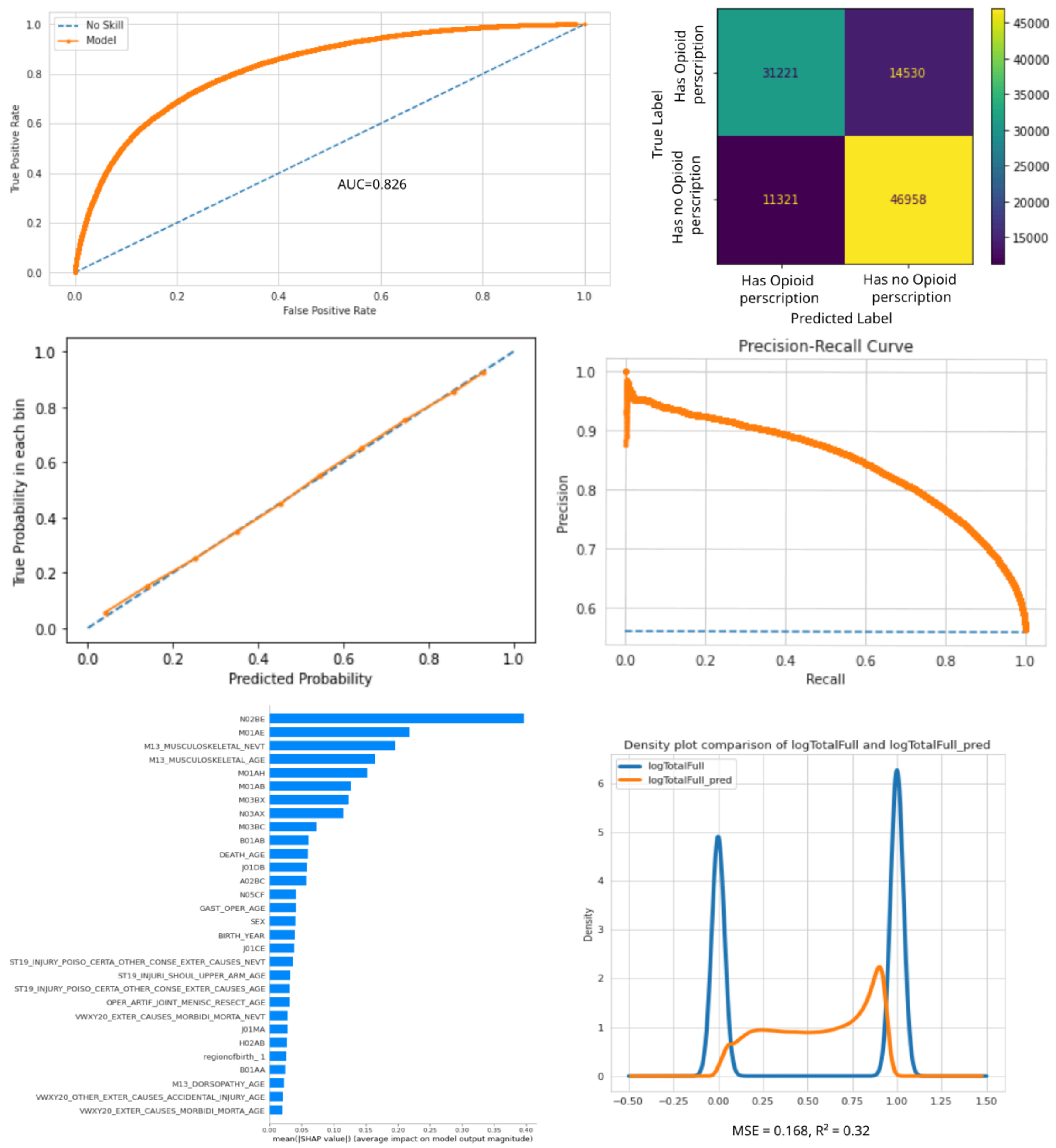


**Supplementary Figure 15 | Performance of the XGBoost model predicting opioid prescription status Model III.**

(a) Receiver operating characteristic (ROC) curve with area under the curve (AUC = 0.825).
(b) Confusion matrix showing 31,221 true positives, 46,958 true negatives, 14,530 false positives, and 11,321 false negatives.
(c) Calibration plot, demonstrating close alignment between predicted and observed probabilities.
(d) Precision-recall curve, with precision >0.6 across most recall values.
(e) Top predictors identified using SHAP values, including musculoskeletal (M13), dorsopathy (M13), and injury-related codes.
(f) Density comparison of observed (blue) and predicted (orange) distributions of binary RxExpPop, with mean squared error (MSE = 0.168) and $R^2$ = 0.32.

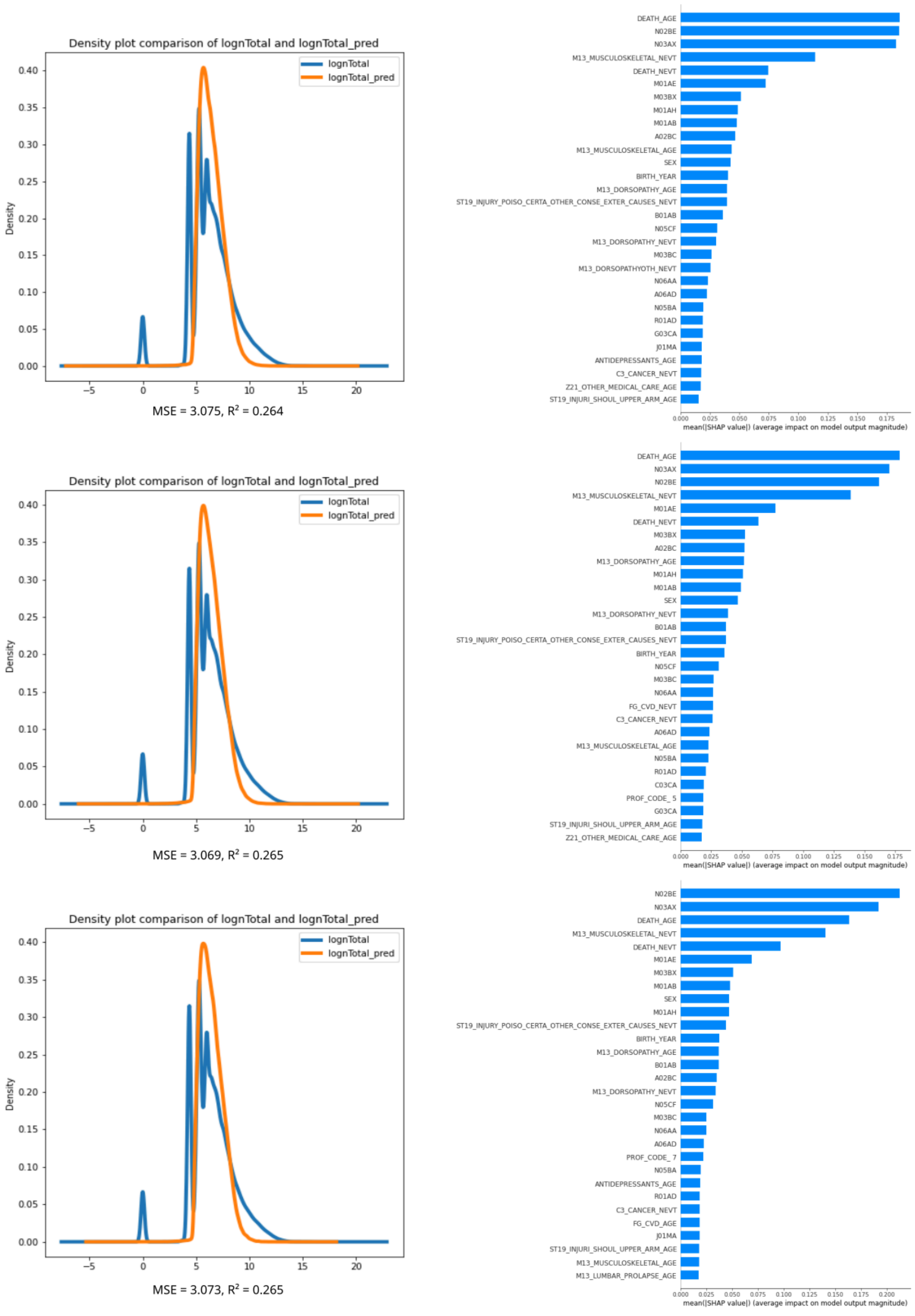


**Supplementary Figure 16 | Performance of three XGBoost models predicting log-transformed opioid dosage (RxDoseUsers).**

(a, c, e) Density plots comparing observed (blue) and predicted (orange) log-transformed opioid dosage across three independently trained models. Model performance was similar, with mean squared error (MSE = 3.075, 3.069, 3.073) and explained variance ($R^2$ = 0.264, 0.265, 0.265), respectively.

(b, d, f) SHAP value plots highlighting top predictors for each model. Consistent predictors across models included age, sex, musculoskeletal (M13) and dorsopathy diagnoses, injury-related codes, and cancer-related codes.

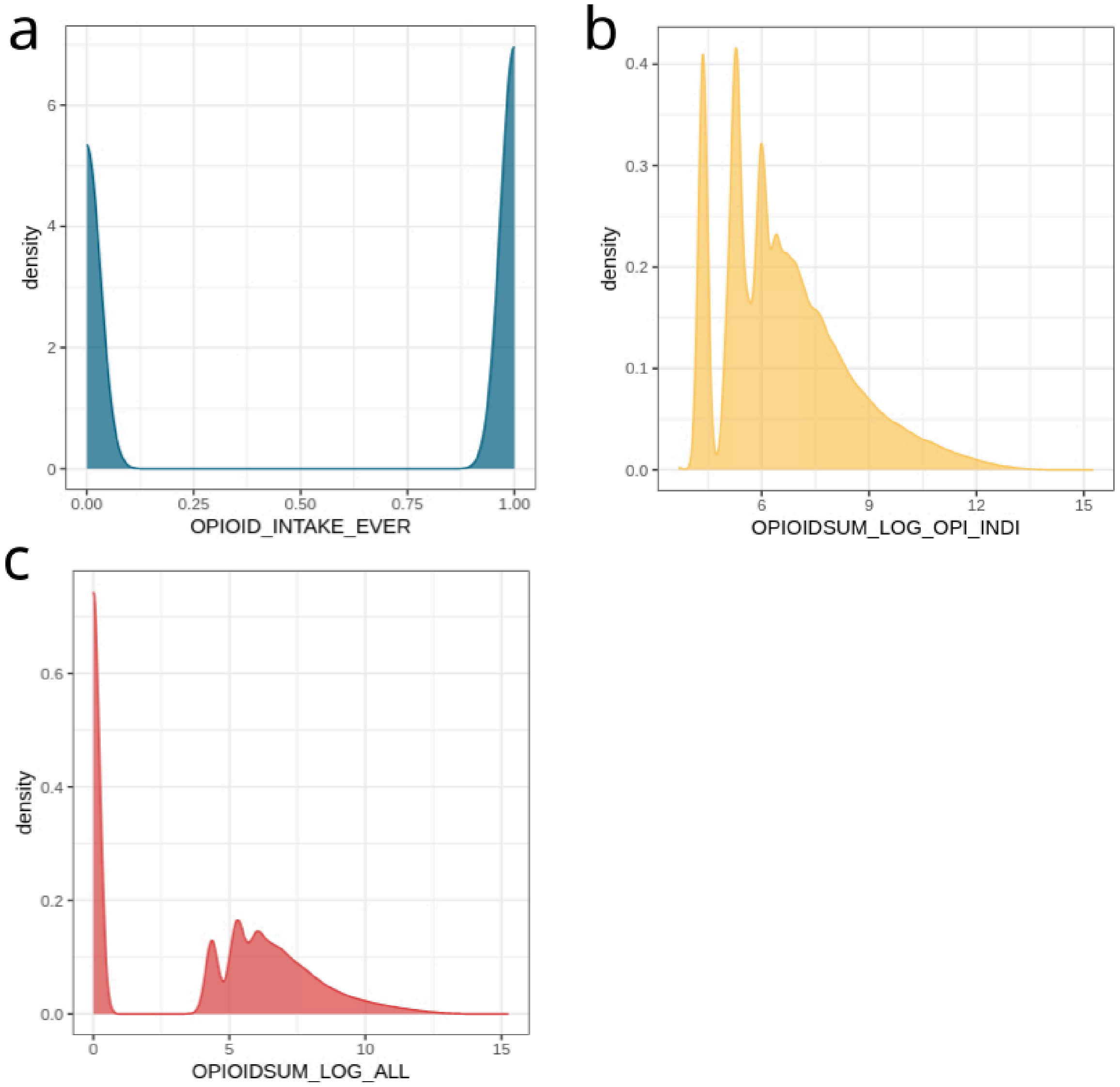


**Supplementary Figure 17 | Density distributions of opioid prescription phenotypes.**

(a) *RxExpPop* (binary ever-prescribed phenotype).
(b) *RxDosePop* (combined dosage plus binary prescription), $log_n$-transformed.
(c) *RxDoseUsers* (dosage among users only), $log_n$-transformed.
